# Altered structural connectome of children with Auditory Processing Disorder: A diffusion MRI study

**DOI:** 10.1101/2022.11.03.22281918

**Authors:** Ashkan Alvand, Abin Kuruvilla-Mathew, Reece P. Roberts, Mangor Pedersen, Ian J. Kirk, Suzanne C. Purdy

## Abstract

Auditory processing disorder (APD) is a listening impairment that some school-aged children may experience as difficulty understanding speech in background noise despite having normal peripheral hearing. Recent resting-state functional magnetic resonance imaging (MRI) has revealed an alteration in regional, but not global, functional brain topology in children with APD. However, little is known about the brain structural organization in APD. We used diffusion MRI data to investigate the structural white matter connectome of 58 children from 8 to 14 years old diagnosed with APD (n=29) and children without hearing complaints (healthy controls, HC; n=29). We investigated the rich-club organization and structural connection differences between APD and HC groups using the network science approach. The APD group showed neither edge-based connectivity differences nor any differences in rich-club organization and connectivity strength (i.e., rich, feeder, local connections) compared to HCs. However, at the regional network level, we observed increased average path length (APL) and betweenness centrality in the right inferior parietal lobule and inferior precentral gyrus, respectively, in children with APD. HCs demonstrated a positive association between APL in the left orbital gyrus and the listening-in-spatialized-noise-sentences task, a measure of auditory processing ability. This correlation was not observed in the APD group. In line with previous functional connectome findings, the current results provide evidence for altered structural networks at a regional level in children with APD, and an association with listening performance, suggesting the involvement of multimodal deficits and a role for structure-function alteration in listening difficulties of children with APD.

## 1. INTRODUCTION

Auditory processing disorder (APD) is a term used to describe children who experience atypical difficulty in understanding speech or other complex auditory stimuli, particularly in the presence of background noise or in quiet, and been diagnosed using a battery of clinical tests (American Speech-Language-Hearing Association, 2005; Dawes et al., 2008; Dillon & Cameron, 2021; Keith et al., 2019; Sharma et al., 2014) It is estimated that globally 5.1% and locally (i.e., in New Zealand) 6.2% of school-aged children have difficulties understanding speech in the classroom despite showing normal hearing sensitivity based on the pure tone audiogram (Hind et al., 2011; Keith et al., 2019; Purdy et al., 2018). This listening difficulty is believed to arise from the complex processing of auditory information in the central auditory nervous system (CANS) in conjunction with other sensory and/or higher-order brain network processing involved in language, hearing, auditory, attention and memory (American Academy of Audiology (AAA), 2010; Dillon & Cameron, 2021; Keith et al., 2019; Moore, 2012; Ponton et al., 1996; Wilson, 2018). APD is heterogeneous and can co-occur with other neurodevelopmental disorders such as attention-deficit/hyperactivity disorder (ADHD), autism spectrum disorder (ASD), dyslexia/reading disorder and/or specific language impairment (SLI) (Dawes et al., 2009; Gokula et al., 2019; Halliday et al., 2017; Mealings & Cameron, 2019; Sharma et al., 2009). It is anticipated that 40 to 56% of children diagnosed with APD also have other comorbid conditions (Ahmmed et al., 2014; Gokula et al., 2019). The overlap of APD symptoms with other sensory or cognitive neurodevelopmental disorders has raised questions about whether APD solely arises from atypical auditory sensory processing (bottom-up approach: related to the ear or CANS) or whether cognitive differences also contribute (top-down approach to cognitive function from multi-modal processing) (Cacace & McFarland, 2013; Dawes & Bishop, 2009, 2010; Dillon & Cameron, 2021; Dillon et al., 2012; Iliadou et al., 2018; McFarland & Cacace, 2014; Moore, 2012, 2018; Moore & Hunter, 2013). It has been suggested that utilizing neuroimaging approaches may help researchers and clinicians to differentiate the neural mechanisms underlying APD (American Academy of Audiology (AAA), 2010; Bartel-Friedrich et al., 2010; Moore & Hunter, 2013; Schmithorst et al., 2011).

In the past few decades, diffusion tensor imaging (DTI), has been widely used to study white matter (WM) microstructural changes in neurodevelopmental disorders (Ameis et al., 2011; Beare et al., 2017; Billeci et al., 2012; Ercan et al., 2016; Sihvonen et al., 2021). Due to its sensitivity to microstructural tissue properties, DTI can be used as a clinical tool to study WM anatomy and the brain’s structural connectome by providing fiber orientation and quantitative diffusion measures (i.e., scalar measures) such as fractional anisotropy (FA - measures fiber density, axonal diameter and myelination of WM), mean diffusivity (MD - measures rotationally invariant magnitude of water diffusion) as well as the axial and radial diffusivity (AD - measures the magnitude of diffusion parallel to fiber tracts, RD - measures the magnitude of diffusion perpendicular to fiber tracts) (Beaulieu, 2002; Soares et al., 2013). These measures have been previously used to study auditory pathways in children with sensory processing disorder (Chang et al., 2014; Owen et al., 2013) and congenital sensorineural hearing loss (Huang et al., 2015; Park et al., 2018). In the APD literature, to our knowledge, two studies have investigated WM microstructure in APD by assessing the association between DTI scalar measures and APD diagnostic test variables (Farah et al., 2014; Schmithorst et al., 2013).

Early research (Schmithorst et al., 2013) investigated whether right or left ear advantage scores on dichotic listening tests (REA/LEA), indicators of hemispheric dominance for auditory processing, language, and learning disorders, can be predicted using DTI and functional MRI techniques. Dichotic listening with speech-related stimuli is typically included in APD test batteries; this task provides information on selective attention and lateralization of brain function within the auditory system. An REA is commonly observed in dichotic listening (Hugdahl, 2011; Moncrieff, 2011). Schmithorst et al.’s research included 24 children diagnosed with (n=10) or without (n=14). During the dichotic listening task, there was greater AD in the sublenticular part of the left internal capsule in the APD group compared to healthy controls (HC). A follow-up DTI study by the same research team was conducted on children with APD (n=12) and HC (n=12) to identify biomarkers of listening difficulties based on WM microstructures (Farah et al., 2014). Correlation analyses based on performance on dichotic listening tasks showed that for participants with APD who had an LEA, there was reduced FA in the bilateral prefrontal cortex and left anterior cingulate and increased MD in the posterior limb of the internal capsule. These results suggest that listening difficulties in children with APD are associated with altered WM microstructure, with sensory and supramodal differences underlying the group differences in auditory processing performance. Although these studies have utilized DTI metrics on children with APD to investigate the relationship between measures of listening difficulties and WM microstructures, no study has yet reported the brain structural connectome in children with APD.

In recent years, network neuroscience has become a promising tool for studying the complex network topology of the brain (Bassett & Sporns, 2017; Bullmore & Sporns, 2009; Sporns, 2011). This method utilizes graph theory by modelling the brain as a network composed of nodes and edges to investigate brain structural and functional connectome (Bullmore & Sporns, 2009). In the structural network, nodes are represented by brain cortical and subcortical regions, and edges are defined by WM streamlines (i.e., fiber) reconstructed by deterministic or probabilistic tractography physically connecting a pair of areas (Fornito et al., 2016d; Rubinov & Sporns, 2010). Within this conceptual framework, brain topological properties such as hub architecture can be studied. Hubs are regions with high levels of connectivity that are spatially distributed but topologically central based on functional measures (i.e., putative brain hubs) (Fornito et al., 2016b; van den Heuvel & Sporns, 2011, 2013).

Hub regions play an important role in global information flow across the brain, which makes them vulnerable spots in the network (Crossley et al., 2014; Fornito et al., 2015; Stam, 2014). Topological measures have been widely used to characterize atypical brain network topology in brain diseases and disorders (Alvand et al., 2022; Crossley et al., 2014; DeSalvo et al., 2014; Fang et al., 2020; W. Li et al., 2016; Lu et al., 2021; Meunier et al., 2009; Roger et al., 2020; Rubinov & Bullmore, 2013; Yuan et al., 2015). Notably, studies have revealed that, in general, the brain hub structure shows densely interconnected and rich organization within hub regions, called the rich-club phenomenon (Pedersen & Omidvarnia, 2016; van den Heuvel & Sporns, 2011). The rich-club is a hierarchical organization where hub regions (i.e., core regions) tend to link more densely among themselves than peripheral regions, providing interregional brain communication and integration and enabling global neural signaling (Colizza et al., 2006; Opsahl et al., 2008; van den Heuvel & Sporns, 2011; Zhou & Mondragon, 2004). The strength of connection in the rich-club organization is categorized as *rich* - the link between cores, *feeder* - the link between core and peripheral, and *local* - the link between peripherals. The term rich club is derived from the analogy in social contexts where highly central individuals (i.e., rich in their connections) create extremely interconnected networks (i.e., through clubs) (van den Heuvel & Sporns, 2011; Zhou & Mondragon, 2004). The rich-club topology can provide important information for integrated communication in brain networks (Colizza et al., 2006; van den Heuvel & Sporns, 2011). Consequently, studying the topological architecture of a rich-club organization could uncover pathological bases for brain diseases (Daianu et al., 2015, 2014; Liu et al., 2021; Lu et al., 2021; Shu et al., 2018; Van Den Heuvel et al., 2013; Xue et al., 2020) and disorders (Cui et al., 2022; Keown et al., 2017; Lou et al., 2021; Ray et al., 2014; B. Wang et al., 2021). A recent connectome-based study of 31 children with sensorineural hearing loss (SNHL) and 31 HC revealed alterations in the rich-club organization in children with SNHL (Cui et al., 2022). The strength of local connections in the SNHL group was significantly higher than in HC. However, there were no differences in rich and feeder connections (Cui et al., 2022). The authors concluded that the alteration in information transmission did not modify the global topology, and the increased local connection may result from insufficient synaptic pruning caused by hearing deprivation (Cui et al., 2022). Thus, investigating the brain hub and rich-club organization of children with APD may advance our understanding of the pathobiology of this neurodevelopmental disorder.

We investigate large-scale WM network organization of children with APD using graph-theoretical analyses. In the present study, we utilized DTI data to construct the brain structural network to explore the brain hub and rich-club architecture of children diagnosed with APD and HC (children without complaints of listening difficulties).

Additionally, we assessed the structural connectivity differences between these two groups (edge-based connectivity). In a recent resting-state functional MRI study (rsfMRI) on children with and without a diagnosis of APD (Alvand et al., 2022), we investigated brain functional hub topology. Our study suggests that functional brain networks in APD were similarly integrated and segregated (i.e., the whole brain averaged network) compared to HC but were significantly different within the default mode network (DMN) in bilateral superior temporal gyrus (STG). Similar to our previous research with functional brain imaging, we hypothesized that the structural connectome is normal in APD on a whole-brain level. Nonetheless, the brain’s white matter may be affected within specific regions involved in auditory and related processing functions.

## 2. MATERIAL AND METHODS

### 2.1 Participants

Sixty-six children aged 8-14 years were recruited for this research as part of previous research (Alvand et al., 2022); 8 participants were excluded from this analysis due to incomplete scans (*n*=4) or head motion (*n*=4). Of the remaining participants, 29 were diagnosed with APD (14 boys, Age=10.89±1.53) and 29 were healthy controls (HC, 14 boys, Age=11.93±1.41). Children diagnosed with APD were recruited from the SoundSkills clinic (https://soundskills.co.nz) in Auckland, New Zealand, based on New Zealand’s standard guidelines for APD test batteries (Keith et al., 2019).

Children in the HC group were recruited via flyers and online advertisements based on the absence of hearing loss or hearing difficulties, neuropsychiatric disorders, or medication affecting the central nervous system. In the APD group, 11 children were also diagnosed with comorbid disorders such as attention-deficit/ hyperactivity disorder (ADD/ADHD, n=2), dyslexia (n=8), and developmental language disorder (DLD, *n*=1). In the HC group, four children were diagnosed with comorbid disorders such as ADHD (n=2), dyslexia (n=1), and autistic spectrum disorder (ASD, n=1), but they were not experiencing any hearing or learning difficulties. These comorbidities were not excluded as they coexist with APD (Dawes & Bishop, 2010; O’Connor, 2012; Sharma et al., 2009). This study was approved by the University of Auckland Human Participants Ethics committee (Date: 18/10/2019, Ref. 023546). Before completing any testing or brain imaging, children and their parents consented to participate in the study. They received financial vouchers to compensate for their participation.

#### Procedure

Our previous study described all pediatric recruitment procedures (Alvand et al., 2022). In summary, children and their parents were invited to attend two individual sessions daily to complete hearing assessments and MRI scans. For the first session, children were tested for hearing acuity, middle ear disease and atypical ipsilateral middle ear muscle reflexes using otoscopy, pure tone air conduction audiometry (PTA), and tympanometry (Roup et al., 1998). All children had a PTA threshold of less than 20 dB HL at octave-interval frequencies from 0.25 to 8 kHz in both ears. Tympanogram results showed static admittance in the range of 0.2 to 1.6 mmho, with peak pressure between -100 and +20 daPa, indicating normal middle ear function. Children were also administered the listening-in-spatialized- noise-sentences (LiSN-S) (Sharon Cameron & Dillon, 2007, 2008), which assesses their ability to hear and remember the target sentences in the presence of competing noise and distraction. More details regarding the LiSN-S sores were described by (Alvand et al., 2022; Besser et al., 2015). Participants were asked to attend an approximately 24 min MRI scan for the second session, including a T1-weighted image (T1w), rsfMRI, and diffusion MRI (dMRI). During T1w and dMRI sequences, participants were asked to stay still and not to move their heads or laugh while watching the movie. Earplugs and headphones were also provided to decrease the scan’s noise.

### 2.2 Data acquisition

All MRI scans are acquired on the same 3T Siemens scanner SKYRA and 20- channel head coil at the Centre for Advanced MRI (CAMRI), the University of Auckland. Diffusion MRI was acquired using single-shot echo-planar imaging (EPI) sequences (TR = 5000 ms, TE = 63.80 ms, FOV: 240 mm, slice thickness: 2.5 mm, voxel size: 2.5mm^3^, Slices: 60, flip angle: 90 degrees, GRAPPA factor: 2, Phase encoding direction: AP, Multi-band acceleration factor: 2). dMRI images were obtained based on 64 diffusion-weighted directions with b-value= 1000 s/mm^2^, and five interspersed scans where b = 0 s/mm^2^. In addition, a single b = 0 s/mm2 was obtained with a reversed-phase encoding direction for susceptibility field estimation. The total duration of the scan was approximately 11 min with 65 volumes. Prior to the dMRI acquisition, a high-resolution structural T1w (4 min; 36 s) was acquired for co-registration and parcellation using a magnetization-prepared rapid acquisition gradient echo (MPRAGE) sequences (1 mm isotropic resolution (FOV 256 mm, 208 sagittal slices in a single slab, TR: 2000 ms, TE: 2.85 ms, Flip angle: 8 degrees, Slice thickness: 1 mm).

### 2.3 Image preprocessing

Initially, all anatomical and diffusion images were converted to NIFTI file sets using dcm2niix version 11/11/2020 (Rorden et al., 2007). Then NIFTI file sets were structured according to the Brain Imaging Data structure (BIDS v1.8.2) (Gorgolewski et al., 2017). The dMRI preprocessing was performed using the QSIprep pipeline version 0.15.3 (Cieslak et al., 2021), based on Nipype version 1.7.0 (Esteban et al., 2022; Gorgolewski et al., 2011). QSIprep is a robust integrative software platform for preprocessing and reconstructing dMRI sampling schemes which leverage metadata in BIDS format to automatically configure suitable processing workflows (Cieslak et al., 2021).

#### Anatomical data preprocessing

The T1w image was corrected for intensity non-uniformity (INU) using N4BiasFieldCorrection (ANTs v2.3.1(Tustison et al., 2010) and used as a T1w-reference throughout the workflow. The T1w-reference was then skull-stripped using antsBrainExtraction.sh (ANTs v2.3.1), using OASIS as the target template. Spatial normalization to the ICBM 152 Nonlinear Asymmetrical template version 2009c (Fonov et al., 2009); RRID: SCR_008796) in the Montreal Neuroimaging Institute space (MNI) was performed through nonlinear registration with antsRegistration (ANTs version 2.3.1, RRID: SCR_004757) (Avants et al., 2008), using brain-extracted versions of both T1w volume and template. Brain tissue segmentation of cerebrospinal fluid (CSF), white matter (WM), and grey matter (GM) were performed on the brain-extracted T1w using FAST (FSL v6.0.5.1:57b01774, RRID: SCR_002823) (Zhang et al., 2001).

#### Diffusion data preprocessing

Any images with a b-value less than 100 s/mm^2^ were treated as a *b = 0* image. MP-PCA denoising, as implemented in MRtrix3’s dwidenoise (Veraart et al., 2016), was applied with a 5-voxel window. After MP-PCA, B1 field inhomogeneity was corrected using dwibiascorrect from MRtrix3 with the N4 algorithm (Tustison et al., 2010). After B1 bias correction, the mean intensity of the DWI series was adjusted so all the mean intensity of the *b = 0* images matched across each DWI scanning sequence. Initial motion correction was performed using only the *b = 0* images. An unbiased *b = 0* template was constructed over 3 iterations of Affine registrations. The SHORELine method was used to estimate head motion in *b > 0* images. This entails leaving out each *b > 0* images and reconstructing the others using the 3D Simple Harmonic Oscillator-based Reconstruction and Estimation (3dSHORE; (Merlet & Deriche, 2013). The signal for the left-out image serves as the registration target. A total of 2 iterations were run using an Affine transform. Model-generated images were transformed into alignment with each *b > 0* image. Both slicewise and whole-brain QC measures (cross-correlation and R^2^) were calculated. A deformation field to correct susceptibility distortions was estimated based on two EPI references with opposing phase-encoding directions, using 3dQwarp (AFNI, (Cox & Hyde, 1997). Based on the estimated susceptibility distortion, an unwarped *b = 0* reference was calculated for a more accurate co-registration with the anatomical reference (see **Fig. 1A**).

**Figure 1.**
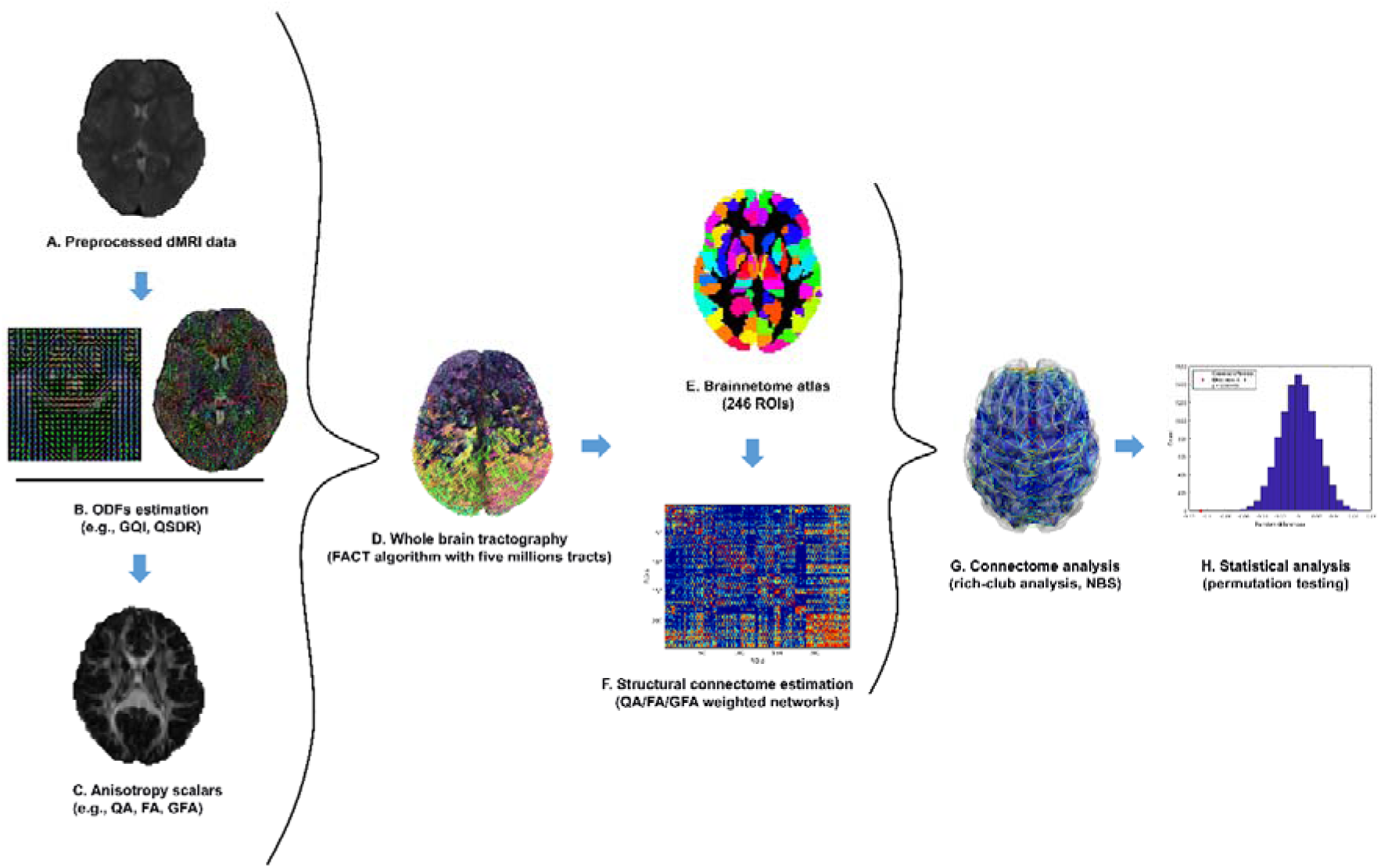
Schematic overview of the study pipeline. (A) diffusion MRI (dMRI) data were preprocessed using the QSIprep pipeline (Cieslak et al., 2021). (B) the output from preprocessing pipeline was then reconstructed using generalized q-space sampling imaging (GQI) and the diffusion orientation distribution functions (ODFs) were recreated in the MNI template space (Q-Space Diffeomorphic Reconstruction, QSDR) (Yeh et al., 2010). (C) quantitative anisotropy (QA), fractional anisotropy (FA) and generalized FA were estimated for individual data. (D) Whole brain tractography was carried out using a deterministic algorithm (fiber assessment by continuous tracking, FACT). (E) brain regions were parcellated into 246 regions of interest (ROIs) using the Brainnetome atlas (Fan et al., 2016) and, (F) structural connectivity was estimated using QA, FA and GFA as network edge weights. (G) Next, network analysis, such as rich-club organization (van den Heuvel & Sporns, 2011) and edge- wise connectivity analysis (i.e., network-based statistics, NBS) (Zalesky, Fornito, & Bullmore, 2010), were carried out. (H) Finally, statistical analyses were performed on individual networks using permutation analysis of linear models (PALM) (Winkler et al., 2014).

The output from QSIprep labeled bylabelledffix *_space-T1w_desc-preproc_dwi.nii.gz* was used for further processing. The implementation of the DSIstudio (https://dsi-studio.labsolver.org/) workflow (i.e., dsi_studio_gqi) in the QSIprep pipeline was utilized to perform diffusion reconstruction, whole-brain tractography, and brain network construction (Cieslak et al., 2021; Yeh et al., 2010). Diffusion orientation distribution functions (ODFs) were reconstructed in template space (i.e., MNI ICBM152) using q-space diffeomorphic reconstruction QSDR (Yeh et al., 2013), which is the MNI version of generalized q-sampling imaging GQI (Yeh et al., 2010) with a ratio of mean diffusion distance of 1.25. The GQI is a model-free method for quantifying the density of water diffusion in various orientations while providing directional and quantitative information regarding crossing fibers (Yeh et al., 2013, 2010) (**Fig. 1B**).

Many internal operations of QSIPrep use Nilearn 0.9.0 (Abraham et al., 2014); RRID: SCR_001362) and Dipy (Garyfallidis et al., 2014). For more details on the pipeline, see the section corresponding to workflows in QSIPrep’s documentation (https://qsiprep.readthedocs.io/en/latest/preprocessing.html).

#### Quality control (QC)

The anatomical and dMRI data were initially inspected for QC assurance using QSIprep’s visual reports (Cieslak et al., 2021). All data were visually assessed for accurate alignment of T1w images and noticeable signal dropouts. The QC of dMRI data was performed using QSIprep’s QC report, as described in Yeh et al. (2019). Based on this QC, image dimensions, image resolutions, DWI count, and b-table were checked for consistency within the dataset. Then, the diffusion image quality metric was evaluated by neighboring DWI correlation (NDC) which summarizes the pairwise spatial correlation between each pair of volumes that sample the closest points in q-space (Cieslak et al., 2021; Yeh et al., 2019). The NDC with lower values represents decreased data quality derived from misalignment between volumes, prominent eddy current artifacts, head motion artifacts, or any head coil issues that might impact the diffusion signals (Yeh et al., 2019). Based on NDC scores, the QC procedure applies the outlier checking function (i.e., 3 median absolute deviations) to label noisy data with *Low-quality outlier* labels (Yeh et al., 2019). For our dataset, the NDC values were high and similar across all dMRI volumes (0.906 ± 0.008). Only four outlier scans were identified based on corrected NDC scores (N_APD_=3, N_HC_=1).

### 2.4 Network construction

Whole-brain fiber tracking for each participant was performed using a deterministic tractography using a modified fiber assessment by continuous tracking (FACT) algorithm (S. Mori et al., 1999; Susumu Mori & van Zijl, 2002). Five million streamlines were reconstructed, and tracts less than 10 mm and greater than 400 mm were eliminated (**Fig. 1D**). For the fiber tracking, the angular threshold of 45 degrees and step size of 0.94 mm were set. The quantitative anisotropy (QA) scalar was then calculated along the path of each reconstructed streamline. The QA is an index of GQI, a measure of anisotropic spins that diffuse along the fiber orientation (Yeh et al., 2010). In other words, QA is the density of anisotropic diffusing water after removing the isotropic components. Along with QA, fractional anisotropy (FA: DTI measure that quantifies the integrity of the interregional white matter connections) and generalized FA (GFA: a measure that quantifies the variation in diffusion ODF by detecting multiple fiber pathways, which results in better estimation of anisotropy, (Tuch, 2004) were also calculated (**Fig. 1C**). Compared to FA and GFA, the QA is less sensitive to the partial volume effect, and it can improve deterministic tractography by filtering noisy fibers and defining track terminations (Yeh et al., 2013). For the construction of the brain network, brain nodes were defined according to Brainnetome parcellation (http://atlas.brainnetome.org/) with 210 cortical and 36 subcortical regions (246 regions), which provides a fine-grained, cross-validated atlas and contains information on both anatomical and functional connections (Fan et al., 2016) (**Fig. 1E**). Network edges were defined where at least one track connected a pair of regions. This connectivity was determined where connecting track passed through the parcellated areas (i.e., connectivity type: *pass* in DSIstudio). Then edge weights were computed based on the mean QA along tracks connecting any pair of regions of interest (ROIs). The network construction resulted in an individual-specific symmetric undirected weighted connectivity matrix with dimensions of *246 x 246*. In addition to QA-weighted networks, supplemental analyses were carried out to assess the robustness of results on edge weights of every pair of regions defined as mean FA (DSIstudio: dti-fa) and mean generalized FA (DSIstudio: gfa) (**Fig. 1F**).

### 2.6 Connectome analysis

Graph measures for each participant were computed on the undirected weighted structural network to investigate microstructural changes in the brain structural topology. Graph theory analysis was carried out using the brain connectivity toolbox (BCT, version 03/03/2019, https://sites.google.com/site/bctnet/) on MATLAB R2019b (https://mathworks.com/) (See **Fig. 1G**).

#### Edge-wise connectivity

The network-based statistic (NBS) approach (Zalesky, Fornito, & Bullmore, 2010) was conducted on an individual’s structural matrices to assess the significance of any connected sub-network (i.e., component) in the set of altered connections. The NBS is a nonparametric statistical method that controls family-wise error (FWE) to identify the largest connected component in the form of alteration. Initially, a primary statistical threshold (*p* < 0.05, uncorrected) was used to identify connected components and their sizes based on a set of supra-threshold links. Second, to test the significance of each identified connected sub-networks, the empirical null distribution of component size was evaluated using a nonparametric permutation test with 10000 randomizations. Afterwards, two-sample t-tests were performed for each pairwise connection linking 246 brain regions to test group differences in structural connectivity in either direction (two-tailed hypothesis test, Initial *t* threshold = 3.9805). Age was controlled as a covariate.

#### Rich-club organization

The rich-club architecture in a network exists when hub nodes are highly connected, more so than expected by chance (Fornito et al., 2016c; van den Heuvel & Sporns, 2011; Zhou & Mondragon, 2004). This tendency of hubs to be highly linked with each other can be defined by calculating the rich-club coefficient. In the present study, the weighted rich-club coefficient, *φ^W^(k)* (BCT function: *rich_club_wu*), was computed on the group-averaged QA-weighted structural network based on the following equations (Opsahl et al., 2008):

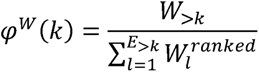

where *k* is the degree of a node, *E*_>k_ denotes the number of edges that exists in the subgraph with degree *> k*, *w*_>k_ is the sum of the weights on edges within the subgraph of nodes with rank greater than *k*, and *w^ranked^* represents a vector of edge weights from highest to lowest across the entire network. *φ^W^(k)* indicates the rich-club effect for each level *k* in the empirical (i.e., observed) network.

Random networks (e.g., Erdős-Rényi model) show characteristics of*φ^W^(k)* by increasing *k* in the network, where nodes with a higher degree are more likely to be interconnected with each other by chance, hence *φ^W^(k)* is normalized with a set from a comparable random network to address this problem (Colizza et al., 2006; Fornito et al., 2016c; McAuley et al., 2007; van den Heuvel & Sporns, 2011). In the current study, 1000 randomized networks were constructed by shuffling the weighted links in the group-averaged network while preserving the degree distribution and sequence of the matrix (BCT function: *randmio_und*, 50 iterations) (Maslov & Sneppen, 2002). The rich-club coefficient was then computed for each random network, 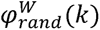 and, for each level *k*, the normalized rich-club coefficient, 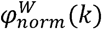, was computed as the ratio between *φ^W^(k)* and average of 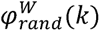 across 1000 networks (Colizza et al., 2006):

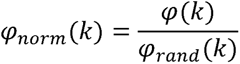

where for each level 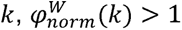 suggests the existence of a rich-club organization in the network (Colizza et al., 2006; Fornito et al., 2016c).

A permutation test was used to evaluate the statistical significance of rich-club organization based on the empirical null distribution of 1000 randomized networks (Bassett & Bullmore, 2009; van den Heuvel & Sporns, 2011). Then, for the range of *k* expressing rich-club organization, 1-tailed permutation tests determine whether *φ^W^(k)* is significantly greater than 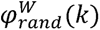 at each level of *k* (one-tailed, *p* < 0.05, 10000 shuffling). This analysis verifies whether any apparent rich-club organization compared to the random topology significantly exceeds 1, indicating the existence of the rich-club organization (Barker et al., 2017; van den Heuvel & Sporns, 2011)

Brain hub regions were identified according to the consensus-based definition of hubs in structural connectivity networks (van den Heuvel et al., 2010), whereby hubs are defined as nodes with high nodal degree (NS, i.e., node strength), high betweenness centrality (BC), low average path length (APL) and low clustering coefficient (CC). More information regarding the definition of the graph measures utilized in this study can be found in Rubinov & Sporns (2010). NS, BC, APL and CC were computed to identify brain hubs based on the group-averaged matrices. Each region was then assigned a score based on the following conditions: (1) the top 20% of regions with the highest NS (*k >* 1 standard deviation above the mean); (2) the top 20% of regions with the highest BC; (3) the bottom 20% of regions with lowest APL; or (4) bottom 20% of regions with lowest CC. Based on these scores (i.e., 0 - 4), ROIs with the highest score (i.e., ≥ *3*) that consistently exist in both groups were selected as brain hubs (Fornito et al., 2016b; van den Heuvel et al., 2010).

Following hub detection, brain regions were classified into core regions (i.e., hub, also known as rich-club regions) and peripheral regions (i.e., non-hub regions). Categorization of brain regions allowed for defining the connections between brain regions into three classes: rich (links between core regions), feeder (links between the core and peripheral regions) and local (links between peripheral areas). For each connectome, the connectivity strength of rich, feeder and local connections was then computed as the sum of all edge weights within each connection class:

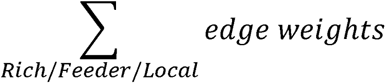

### 2.7 Statistical analysis

Differences in group characteristics (i.e., age, gender, handedness) and LiSN-S variables were tested in the Statistical Package for the Social Sciences (SPSS v28.0). A chi-square test was used to examine gender and handedness effects. Two-sample t-test analyses were employed to assess group differences in age and LiSN-S measures.

#### Group differences

To investigate between-group differences in the rich-club and brain hub organizations, a permutation analysis was conducted for each individual’s brain network using permutation analysis of linear models software (PALM) (Winkler et al., 2014). Two-sample t-tests were used assuming unequal variances for graph metrics of rich-club coefficients, normalized rich-club coefficients and network hub measures (APL, BC, CC, NS) using 20000 randomizations. The impact of age was controlled for as a nuisance covariate (demeaned). To control for the effect of multiple comparisons, all *p* values obtained from graph analysis were corrected across ROIs (*k* level for the case of rich-club metrics) and all measures (i.e., *φ^W^(k)*, 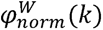, APL, BC, CC, NS) using Bonferroni correction (*p < 0.05*, PALM function: - corrmod). Between-group comparisons were also carried out to assess the differences in three classes of connectivity strength (i.e., rich, feeder, and local). A Bonferroni correction was applied to correct multiple comparisons, with the significance set at *p* < 0.05.

To investigate the relationship between the network parameters that showed significant between-group differences and LiSN-S variables (z-scored), partial correlations were computed using PALM (20000 permutations) (Winkler et al., 2014), for all participants while controlling the effect of age. All *p* values were corrected for the multiple comparisons problem using Bonferroni correction across ROIs and network measures (PALM: -corrmod, *p < 0.05*) (See **Fig. 1H**).

## 3. RESULTS

### 3.1 Demographics

**Table 1** summarizes the demographic characteristics and behavioral measures of APD and HC participants. Age differed between groups (HC *>* APD, *p* < 0.05), but there were no other significant differences in the demographic and LiSNS-S variables. Because of the between-group differences in age, this was included as a nuisance regressor in the statistical analyses. Apart from a statistical trend (*p*= 0.051), the groups did not differ in LiSN-S performance; this test is a part of a wider test battery used to diagnose APD in children. The distribution of LiSN-S variables for all participants is shown in Fig. S1.

**Table 1.** Group demographics

|  | APD (29) | HC (29) | Test Statistic | *p* value |
| --- | --- | --- | --- | --- |
| **Age (years)** | 10.90±1.532 | 11.94±1.416 | 2.693<sup>a</sup> | 0.009 |
| **Gender (male/female)** | 14/15 | 14/15 | 0.000<sup>b</sup> | 1.000 |
| **Handedness (right/left)** | 24/5 | 27/2 | 1.505<sup>b</sup> | 0.220 |
| **LiSN-S (total/missing)** | (27/2) | (28/1) |  |  |
| **Total advantage** | 0.39±1.114 | 0.52±0.991 | -0.273<sup>a</sup> | 0.652 |
| **Spatial advantage** | -0.31±1.525 | 0.24±1.054 | 1.542<sup>a</sup> | 0.129 |
| **Talker advantage** | -0.78±0.966 | -0.30±0.817 | 1.999<sup>a</sup> | 0.051 |
| **High cue** | 0.13±1.183 | 0.50±0.890 | 1.315<sup>a</sup> | 0.194 |
| **Low cue** | -0.33±1.089 | 0.04±0.997 | 1.285<sup>a</sup> | 0.204 |

**Note:** Data are presented as mean ± standard deviation. (a) two-sample t-tests, (b) Pearson chi-square test. HC - healthy control, APD - auditory processing disorder, LiSN-S - Listening in Spatialized Noise-Sentences Test.

### 3.2 Whole-brain structural connectivity is unaffected in APD

The NBS analysis showed no significant differences in individual networks based on the QA-weighted structural connectivity. Further validation analysis based on FA-weighted and GFA-weighted networks also indicated no significant differences in structural connectivity between groups.

### 3.3 Rich-club organization is the same across groups

**Figure 1** illustrates the rich-club organization found in the structural connectome of the APD and HC groups based on the group-averaged QA-weighted network. For both groups, the existence of the rich-club organization (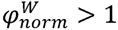) was observed over the range of degrees from *k = 43 - 179*. Consistently, for both groups, the weighted rich-club coefficient of the empirical network was significantly higher than the weighted rich-club coefficient of random networks, based on the range of *k*, indicating robust rich-club organization in the structural network (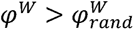, *p* < 0.05). Results from group comparisons based on 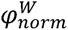 and *φ^W^*showed no significant differences between APD and HC individuals across *k* level (node degree).

Results from hub detection revealed 23 hub regions (i.e., core regions or rich-club regions) for both APD and HC groups. These rich-club regions included the following ROIs according to the Brainnetome atlas: left fusiform gyrus, left precuneus, bilateral parietooccipital sulcus, bilateral hippocampus, bilateral basal ganglia, and bilateral thalamus confirming the results of a previous report (van den Heuvel & Sporns, 2011). More detailed information about hub regions can be found in **Table 2**. After revealing core (hub) regions, peripheral areas were identified, and the connection strengths of rich, feeder, and local were compared between groups. No significant differences between groups were found in the connectivity strength of either rich, feeder, or local connections (**Fig. 2B**).

**Figure 2.**
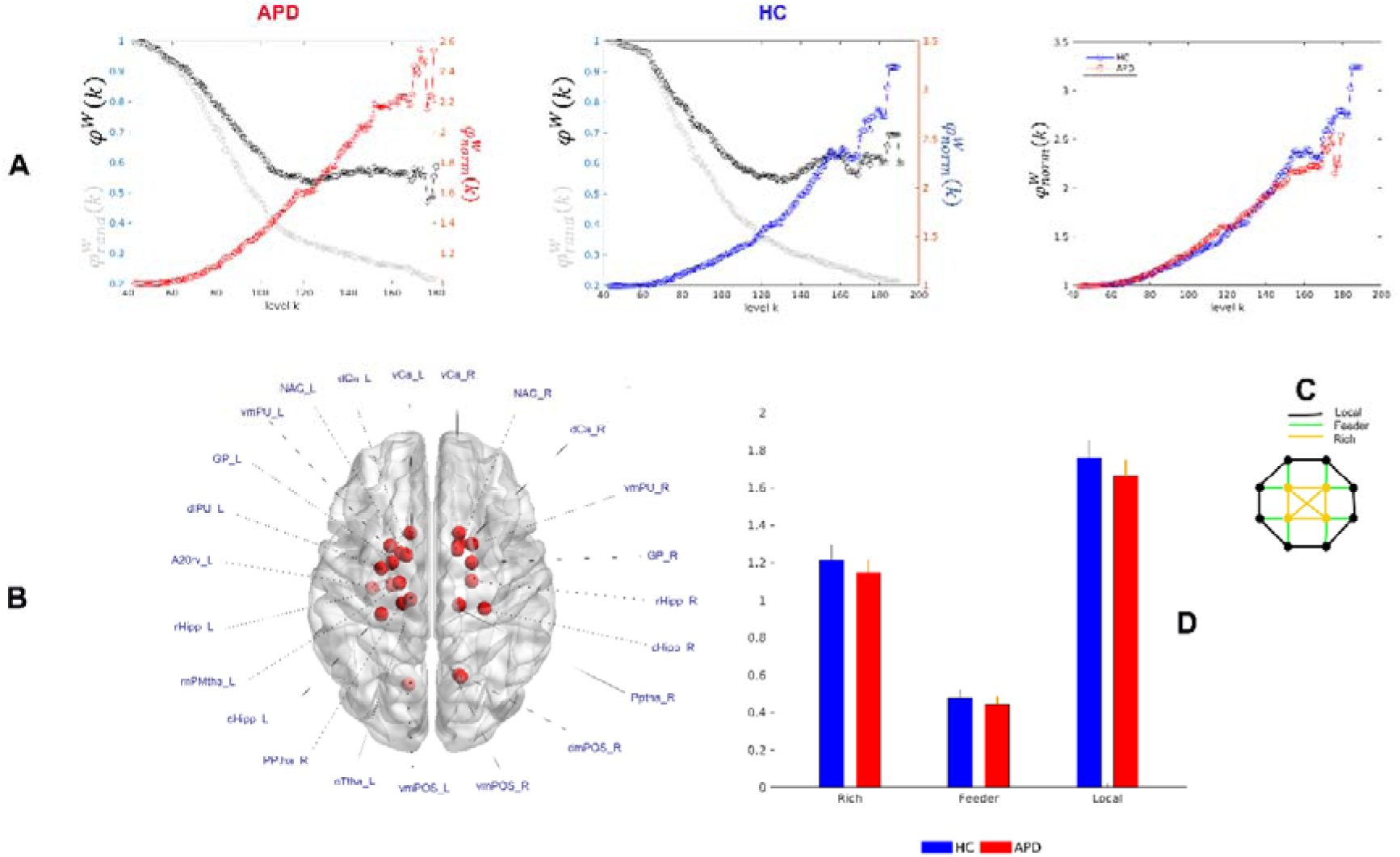
Rich-club organization in children with APD and HC. (A) The empirical rich-club coefficient (black, φ^W^(k)), mean rich-club coefficient of 1000 randomized networks (grey, 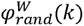), and normalized rich-club coefficient (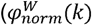) are shown for the group-averaged networks (QA- weighted). Each symbol ‘o’ represents a normalized/rich-club coefficient at each level k. APD and HC groups showed rich-club organizations where 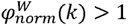. Group comparisons based on individual and group-averaged networks showed no significant differences in the normalized rich-club coefficient. (B) Twenty-three brain hub regions (red balls) were identified for both APD and HC groups based on the frequency of an ROI in four hub measures of NS, BC, APL, and CC. These hub / rich-club regions were found in the left fusiform gyrus, left precuneus, bilateral parietooccipital sulcus, bilateral hippocampus, bilateral basal ganglia, and bilateral thalamus. Brain hub regions were visualized in BrainNet Viewer (Xia et al., 2013). See **Table 2** for more information regarding brain hubs’ labels. (C) The schematic view of rich connections (red) linking rich-club members, feeder connections (blue) linking hub regions to non-hub regions, and local connections (black) linking non- hub regions. (D) Group comparison based on individual’s rich, feeder and local connections indicated no considerable differences between APD and HC groups. The error bars represent a 95% confidence interval of the connection strength in each group. HC - healthy control, APD - auditory processing disorder, NS - nodal strength, BC - betweenness centrality, APL - average path length, and CC - clustering coefficient.

**Table 2.**
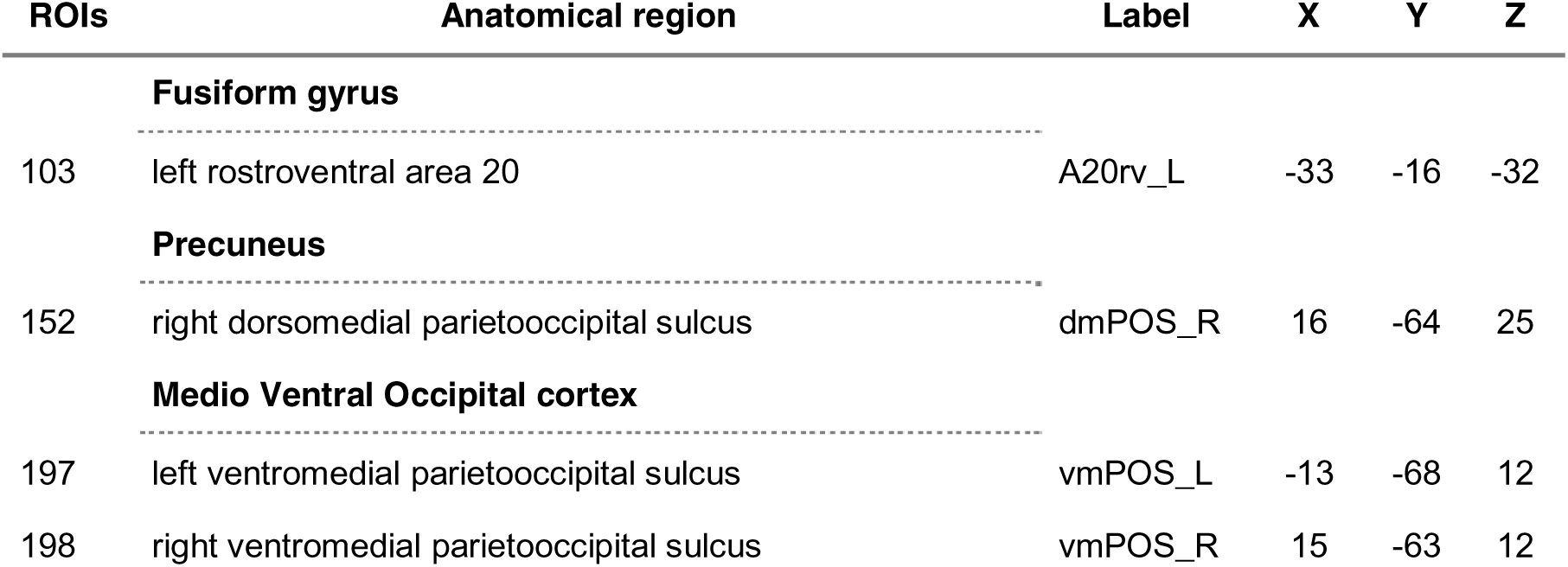
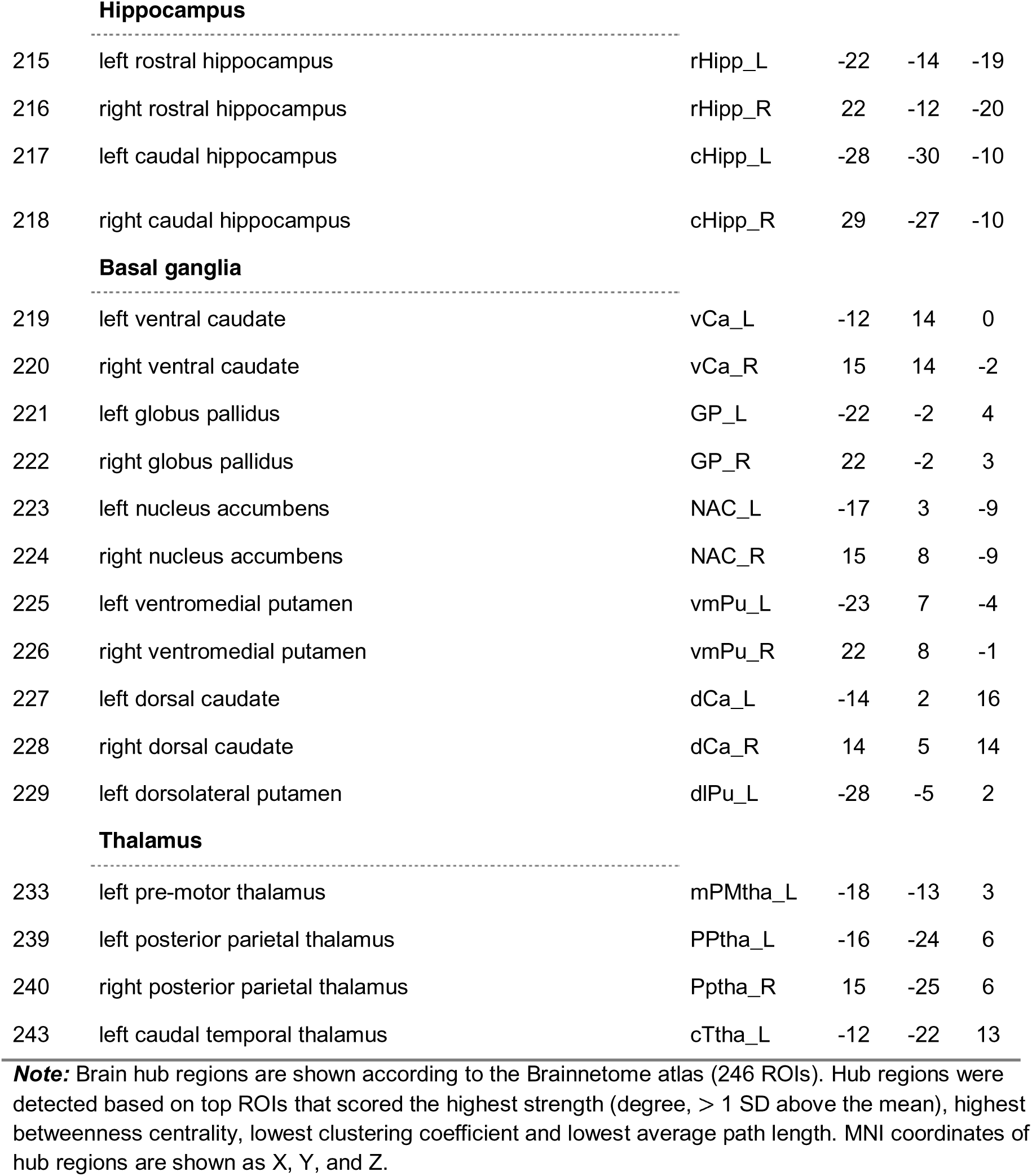
Brain hub regions in APD and HC groups

### 3.4 Nodal differences between APD and HC

**Figure 3** illustrates nodal differences in hub measures between APD and HC groups. Group comparison showed a significant increase in APL measure for the APD individuals (APD *>* HC, *p = 0.0097*, Bonferroni corrected) in the right rostroventral inferior parietal lobule (IPL, label= A39rv_R, ROI #144). The analysis also demonstrated significant between-group differences in BC measure (APD *>* HC, *p = 0.0398*, Bonferroni corrected) in the right ventrolateral precentral gyrus/inferior precentral gyrus (IPG, label= A6cvl_R, ROI #64). No significant group differences were found in CC and NS measures. Validation analysis based on FA-weighted and GFA-weighted networks has also shown group differences in ROI #64 (based on APL measure) and ROI #144 (Based on BC measure); however, they did not pass the multiple comparison correction (supplementary **Table S1**).

**Figure 3.**
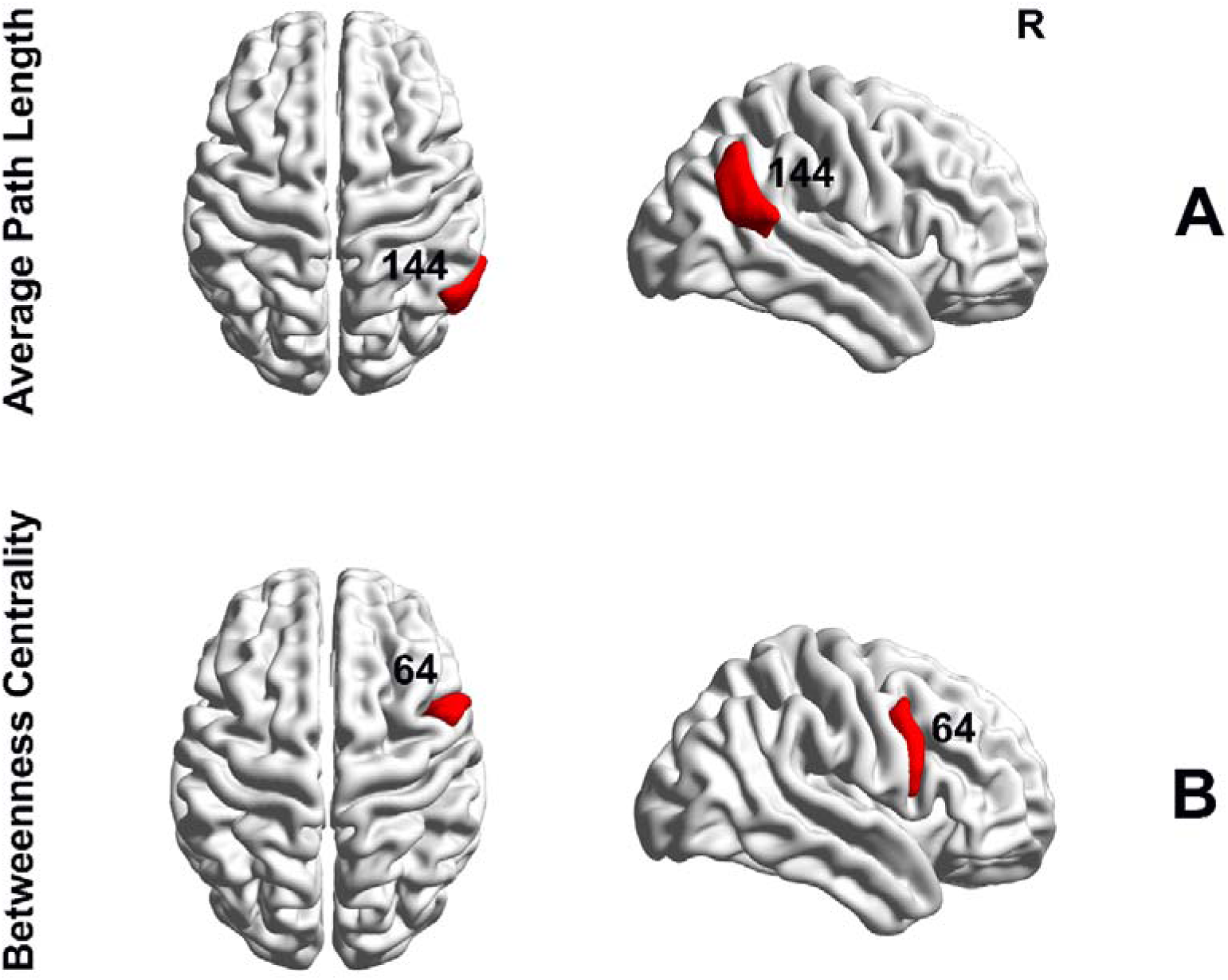
Group differences in nodal hub measures of BC and APL based on Brainnetome parcellation. (A) Significant group differences in the right rostroventral IPL (ROI #144) where APD showed an increase in APL (p = 0.0097, Bonferroni corrected). (B) Significant group differences were found based on BC measure in the right IPG (ROI #64, APD > HC, p = 0.0398, Bonferroni corrected). Brain surfaces in sagittal and axial views were constructed in BrainNet Viewer (Xia et al., 2013). APL - average path length, BC - betweenness centrality, HC - healthy control, APD - auditory processing disorder, R - right hemisphere, IPL - inferior parietal lobule.

### 3.5 Relationship between network metrics and LiSN-S variables

**Figure 4** demonstrates the association between the LiSN-S spatial advantage scores and the network measure of APL. The result indicated significant positive correlations between the spatial advantage and APL in HC groups in the left lateral orbital gyrus (OrG, ROI #51, Pearson r = 0.6216, p < 0.02, Bonferroni corrected). No significant correlation was found between the APD group and the network measure of APL.

**Figure 4.**
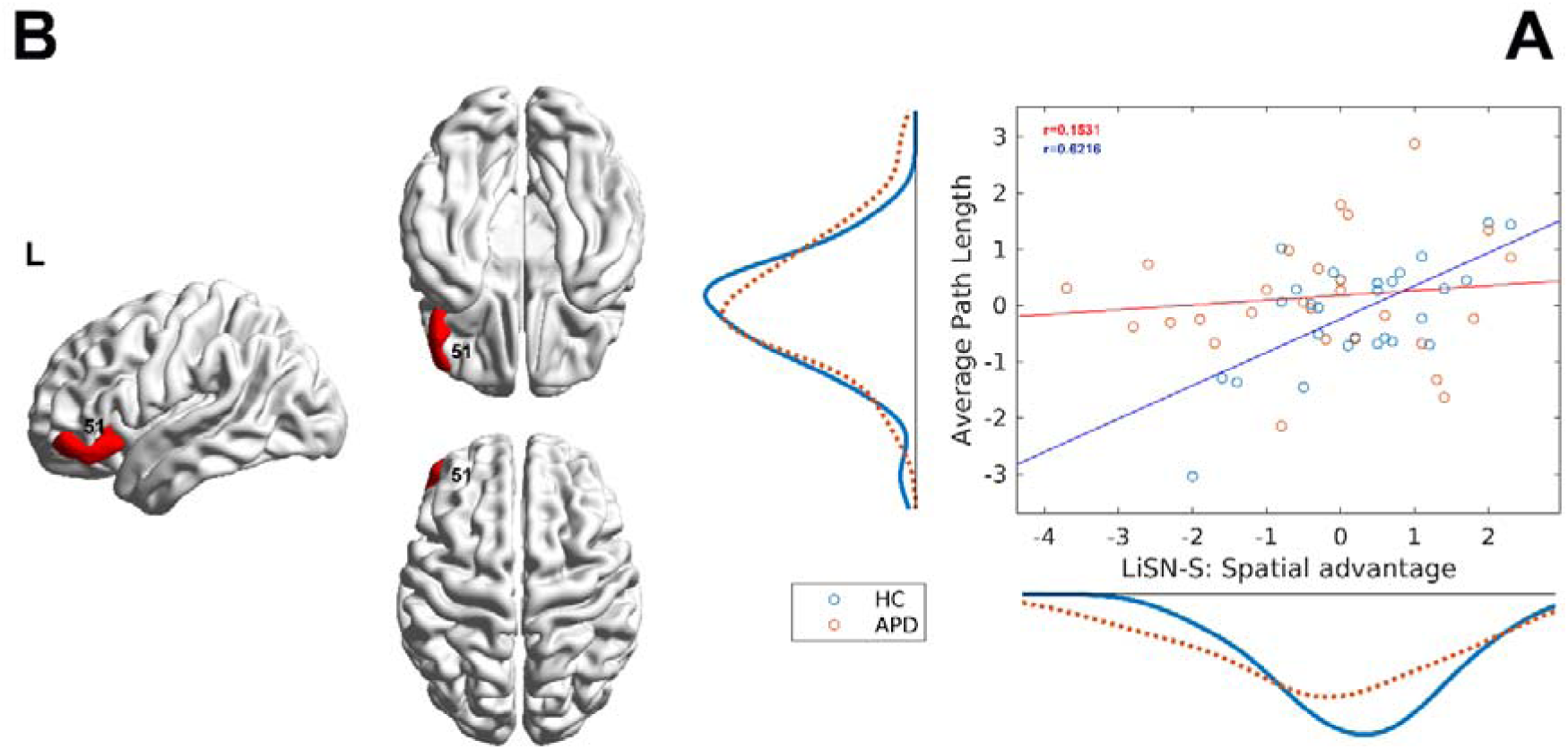
Relationship between LiSN-S variable and the network metric. (A) The scatter plot shows the association between LiSN-S variable spatial advantage (z-scored) and the network metric average path length (APL, z-scored) for both APD (red) and HC (blue) groups in the ROI #51. The marginal distribution of APL (left) and spatial advantage (bottom) are also shown along axes for both groups. The solid lines represent the fitted line. The plot shows a significant positive association for the HC group in the left lateral OrG (p < 0.05, Bonferroni corrected). (B) This association is illustrated in brain surfaces in sagittal and axial views constructed by BrainNet Viewer (Xia et al., 2013). L – left hemisphere, ROI – region of interest, APD – auditory processing disorder, HC – healthy control.

## 4. DISCUSSION

In the present study, we examined brain WM connectome in children diagnosed with and without APD between 8 to 14 years old. This, to our knowledge, is the first study that investigated rich-club organizations using diffusion-based connectivity and a complex network science approach. In line with our hypothesis, the current findings suggest there is a similar global WM structural connectome between APD and HC in terms of rich-club organization (i.e., hub topological structure), the strength of connectivity (i.e., rich, feeder, local) and edge-based connectivity. However, our regional findings (nodal measures of APL and BC) showed significant between-group differences in the right IPL and the right IPG. Additionally, the correlation analysis revealed positive associations between the APL metric and behavioral measure of spatial advantage in the left OrG for HC participants. These findings extend our understanding of the neuropathological mechanisms underlying APD from a perspective of the structural connectome.

### 4.1 Rich-club organization is intact in APD

Rich-club organization is a key characteristic of brain networks, and its existence has not only been found during human brain development (Sa de Almeida et al., 2021; van den Heuvel & Sporns, 2011), but also in numerous neurological diseases (Cao et al., 2014; Collin, Kahn, et al., 2014; Cui et al., 2022; D. Li et al., 2020; Liu et al., 2021; Peng et al., 2021; Ray et al., 2014; Van Den Heuvel et al., 2013). In our study, the rich-club network of densely interconnected hubs was observed for both APD and HC groups in the left fusiform gyrus, left precuneus, bilateral parietooccipital sulcus, bilateral hippocampus, bilateral basal ganglia (i.e., caudate, globus pallidus, putamen, nucleolus accumbens), and bilateral thalamus. The observed rich-club regions were largely consistent with previous reports on hearing-related research by (Cui et al., 2022) on the SNHL participants and other structural brain network studies (Collin, Kahn, et al., 2014; van den Heuvel & Sporns, 2011; Van Den Heuvel et al., 2013). These results suggest the existence of a robust, densely interconnected rich-club in APD connectome. This similarity in the rich-club organization could potentially reflect the intact brain WM structures in APD due to its subtle behavioral differences compared to the HC group. Additionally, identified rich-club regions from the present study form the components of the default mode network (DMN), such as the precuneus/posterior cingulate cortex (Hagmann et al., 2008; Raichle et al., 2001; van den Heuvel & Sporns, 2011) which has been previously reported to has an important role in between module connectivity (van den Heuvel & Sporns, 2011) and contribute in neurocognitive functions (e.g., memory and attention) (Castellanos et al., 2008; Delano-Wood et al., 2012). Previously, the DMN regions, such as the superior temporal gyrus (Alvand et al., 2022; Stewart et al., 2022), and posterior cingulate cortex/precuneus (Pluta et al., 2014) in the functional connectivity studies on children with APD, have been reported to be disrupted indicating the role of functional changes in the brain network.

The connection between the rich-club structure is hypothesised to be a foundation for high-level information transmission (Collin, Sporns, et al., 2014; van den Heuvel & Sporns, 2011; Van Den Heuvel et al., 2013). These dense connections enable the brain to process signals in scattered distributed modular structures and integrate the processed information across all modules through long-distance connections between hub regions (van den Heuvel & Sporns, 2011). Thus, alteration in rich-club connections reflects the brain’s global communications impairment (Stam, 2014; van den Heuvel & Sporns, 2011). Abnormalities in rich (i.e., hub-hub), feeder (i.e, hub- nonhub), and local (i.e., nonhub-nonhub) connections have been reported previously in individuals with neurological disorders along with alteration in local topological metrics (Cui et al., 2022; Lou et al., 2021; Ray et al., 2014; Shu et al., 2018; B. Wang et al., 2021; Y. Wang et al., 2019). In contrast, our results did not show significant between-group differences while comparing the normalized rich-club coefficient (i.e., the measure of rich-club organization) and the strength of connectivity (i.e., rich, feeder and local connections). Still, they indicated group differences in brain networks’ local properties (i.e., APL and BC). This was inconsistent with the results from the recent hearing-related study by (Cui et al., 2022), which reported an increase in local connection on SNHL and no differences in nodal topological measures. This could suggest that our results rely on differences in the topological arrangement of connections and their weights to the rich-club regions rather than the differences in the strength of such a connection (Baldi et al., 2022). Hence, these findings could indicate that the information transmission between structural regions was not significantly changed between both groups. Children with APD have a similar structural connectome compared to the HC group.

### 4.2 Alterations in Regional network in APD

At the local topology level, we found a trend of increase in the nodal measure of APL in the IPL region situated in the DMN for the APD group (ROI #144, **Fig. 3A**). APL is the average number of steps along the shortest path for every possible pair of nodes (Rubinov & Sporns, 2010). APL measures information efficiency, and higher APL suggests less efficiency in information flow (Fornito et al., 2016a). Also, the increase of APL previously in DTI studies on WM connectome was attributed to the degeneration of fiber bundles and disconnection in information transmission (Bai et al., 2012). Furthermore, the meta-analytical relationship between cognitive terms and the intraparietal lobule has indicated an association with the theory of mind, internally-oriented thoughts and autobiographical memory (Yarkoni et al., 2011). This region is located in the DMN, which is involved in self-referential processing, higher-order cognitive functioning and self-emotion regulation (Buckner et al., 2008; Kaiser et al., 2015; Raichle et al., 2001). The aberrant functional connectivity within the DMN has been reported in hearing loss (Schmidt et al., 2013) and APD studies (Alvand et al., 2022; Pluta et al., 2014). Also, this result is in line with our previous fMRI study on children with APD, where between-group comparisons based on participation coefficient metric indicated differences in the right IPL (ROI #298, Schaefer atlas) (Alvand et al., 2022). There have been a few other research in auditory-hearing literature that reported the intraparietal lobule role in the DMN. For instance, an fMRI study on young adults with sensorineural hearing loss confirmed the relationship of IPL with DMN and suggested its role in linguistic thinking (Z. Li et al., 2015). Studies on individuals with unilateral hearing loss (UHL) have also reported increased functional connectivity in the UHL group, where IPL may contribute to the remodeling of the sensory system (Xie et al., 2019). Yang et al. (2014) research on UHL has also reported that IPL, an important region in the DMN, could be susceptible to chronic auditory deprivation.

Furthermore, our results also showed a trend of increase in the BC metric for the APD group in the IPG region located in the executive control network (ECN, ROI #64) (**Fig. 3B**). The BC metric measures the shortest paths that pass through a node (Rubinov & Sporns, 2010). Nodes with a high score of BC participate in numerous shortest paths, meaning higher BC indicates greater influence and control in information transmission (Fornito et al., 2016b; Rubinov & Sporns, 2010). A DTI research on WM connectome reported that an increase of BC in a brain region indicates that the WM network associated with that region is highly affected by shorter/local pathways (Torgerson et al., 2015). Thus, our result based on BC could suggest that the IPG has a critical and central role in the structural connectome of the APD group. Also, meta-analytical correlation showed that IPG is associated with working memory, demands and tasks (Yarkoni et al., 2011). This area, ROI #64, is located in the ECN, which is considered responsible for executive functions that regulate cognitive processes such as working memory, problem solving, planning and reasoning (Qin et al., 2015). The relationship between IFG and working memory was previously reported by (Kambara et al., 2017) study on patients with epilepsy during auditory verbal working memory tasks, which indicated the role of IPG in the initial maintenance of memory cues (Nakai et al., 2019). Studies on hearing loss also reported the increased activity of the right IPG in individuals with longer hearing aids experience suggesting its involvement in speech processing outside of the core speech processing network (Vogelzang et al., 2021). Consistent with the mentioned research, another study on a normal adult with lipreading reported the involvement of IPG in the motor theory of speech, where speech processing is related to the activation of the premotor region (Ruytjens et al., 2006).

On a different note, studies have shown that the DMN is an antagonist of ECN and deactivates during the activation of ECN (Barkhof et al., 2014). Neuroimaging studies on bipolar disorder investigated the alteration in functional connectivity within and between DMN and ECN and suggested dysregulation in cognitive processing (Goya-Maldonado et al., 2016; Y. Wang et al., 2018). The imbalance between these two networks has also been reported could be potentially associated with the self-focus condition and rumination, indicating an inability to reallocate neural resources for effective down-regulation of self-referential thoughts (Belleau et al., 2015; Kaiser et al., 2015).

Additionally, our correlation analysis revealed a positive relationship between APL (i.e., network efficiently) measure and LiSN-S variable, spatial advantage, in the left lateral OrG for the HC group, while no correlation was found for APD group (ROI #51, **Fig. 4A, B**). The left lateral OrG is located in the prefrontal cortex and has been previously associated with semantics, language comprehension, sentence comprehension and language network (Billingsley et al., 2001; Ferstl et al., 2008, 2005; Frost et al., 1999; Gitelman et al., 2005). The spatial advantage is part of the APD diagnosis battery test (Keith et al., 2019) which reflects the individual’s ability to use spatial cues for distinguishing the talker’s directions in the presence of distractions (Cameron et al., 2006a, 2006b, 2011). The link between spatial processing of language comprehension and the left OrG has been previously shown in neuroimaging research (Chow et al., 2014; Rocca et al., 2020; Santesso et al., 2008). Thus, the correlation between spatial advantage and the left OrG APL suggests a greater ability of HC individuals to localize and comprehend the talker’s voice in the language network compared to the APD group.

Together our findings relative to alteration in the regional brain networks in the DMN and ECN could suggest the notion of adaptive neural response in pathological perturbation, such as neural compensatory and degeneracy mechanisms in APD individuals (Fornito et al., 2015). Fornito and colleagues (2015) explained how effectively our brain could use adaptive behavior to maintain its performance during neural insult. They specified that the elevation in functional activity in neurological disorders is commonly attributed to the compensatory mechanisms of the neural system, which can last for an extended period of time with a high degree of preserving behaviors (Fornito et al., 2015). This statement is in line with our results based on structural networks, which are stated in this section and our recent functional research on children with APD, where we showed functional regional network differences in bilateral regions (e.g., superior temporal gyrus and temporo-occipital cortices) (Alvand et al., 2022). Fornito et al. (2015) further explained degeneracy as a complementary mechanism in addition to compensatory.

Degeneracy is defined as the capability of anatomically distinct regions in the brain to execute similar functions and is apparent at multiple levels (e.g., large-scale networks) (Fornito et al., 2015; Tononi et al., 1999). For example, it was noted that during any task, multiple neural networks activate in a parallel and redundant manner to support functional performance. Any possible failure in one system can be replaced by other backup systems (Noppeney et al., 2004). This is aligned with our current results regarding the increase in the structural brain regions located in two segregated functional networks (i.e., DMN and ECN), suggesting the neural basis of cognitive reserve (i.e., ability to engage alternative compensatory mechanisms to encounter behavioral changes) (Barulli & Stern, 2013; Fornito et al., 2015; Valenzuela et al., 2007). Thus, our findings, combined with previous results, align with our recent fMRI research that concluded the involvement of memory and cognitive functioning in APD’s brain network (Alvand et al., 2022) and it could suggest the role of multimodal deficits and structure-function alteration in listening difficulties.

### 4.3 Limitations and future directions

This study has several limitations. First, despite reporting results based on the effective sample size, a larger cohort of participants should be recruited in the future to validate the findings. Second, the results from this study were reported based on Brainnetome structural parcellation (Fan et al., 2016). However, studies showed that the semi-arbitrary brain parcellation selection could affect the results (Andellini et al., 2015; Termenon et al., 2016; Zalesky, Fornito, Harding, et al., 2010). Thus, future research could employ parcellation schemes with a similar number of parcels (i.e., nodes) to mitigate this potential issue. Third, the inherent restrictions of the dMRI sequences and deterministic tractography algorithm would urge us to interpret the results carefully. Using deterministic tractography due to its limitation in resolving crossing, converging, or diverging streamlines (i.e., fiber) and producing inaccurate connectivity matrices (i.e., false positive or false negative fibers) could have influenced the results (Maier-Hein et al., 2017). Although probabilistic tractography could potentially prevent the issue of fiber crossing, the problem of false positive fibers would remain a potential issue (Zalesky et al., 2016). A possible solution to the mentioned limitations could be utilizing advanced dMRI sequences for analyzing diffusion data by fixel-based analysis (FBA, Dhollander et al., 2021). The FBA method allows the construction of fiber population within a voxel (similar to the voxel-based analysis for rsfMRI data) in the presence of complex fiber arrangements, which is considered beneficial compared to the traditional voxel averaged-approach (e.g., DTI) (Dhollander et al., 2021; Raffelt et al., 2015, 2017; Tournier et al., 2019). Using the FBA method, different diffusion indices, as opposed to DTI/GQI method, can be obtained, such as fiber density (FD: microstructural changes in local loss of intra-axonal volume), fiber cross-section (FC: an estimate of macrostructural changes in the diameter of fiber bundle) and fiber density/fiber cross-section (FDC: the combination of FD and FC) (Raffelt et al., 2015, 2017). Future studies on the APD population can use the FBA approach to allow us to expand the understanding of WM connectome’s microstructural and macrostructural properties. Lastly, as discussed in our previous rsfMRI study (Alvand et al., 2022), the heterogeneity of our sample size could have affected the results. We recommend future research employ a longitudinal approach to investigate the developing brain of children with APD until their maturation. This is because the structural connections between brain regions are continuously growing, and exploring the rich-club organization of these changes would give us better insight regarding APD’s brain (Oldham et al., 2021).

## 5. CONCLUSIONS

In summary, our study provided evidence of undisrupted whole-brain WM topological organization and abnormality in the regional structural network located in the right IPL (based on APL metric) and IPG (based on BC metric) within DMN and ECN, respectively. These brain regions are associated with cognitive functioning, such as the theory of mind, autobiographical memory, and working memory. Additionally, correlation analysis with behavioral measures showed a positive association in the left amygdala for all the participants indicating the role of speech processing in this region. Our findings could suggest the involvement of multimodal deficits and a role for structure-function alteration in listening difficulties, providing a new perspective for understanding the pathological mechanisms of APD.

## Data Availability

The data that support the findings of this study are available from the corresponding author upon reasonable request.

## ACKNOWLEDGEMENTS

We would like to thank Soundskill clinic in Auckland for assisting us in participant recruitment and also all the participants and their parents for participating in our study during the COVID- 19 global pandemic. We also thank and acknowledge CAMRI and MRI technicians for facilitating and supporting us in conducting these scans. Further, we would like to thank Eisdell Moore Centre for funding our research.

## Credit author statement

**Ashkan Alvand**: Conceptualization, Methodology, Investigation, Data curation, Software, Formal analysis, Visualization, Writing - original draft, Writing - review & editing. **Abin Kuruvilla-Mathew**: Conceptualization, Methodology, Investigation, Funding acquisition, Supervision, Writing - review & editing. **Reece R. Roberts**: Methodology, Supervision, Writing - review & editing. **Mangor Pedersen**: Methodology, Validation, Writing - review & editing. **Ian J. Kirk**: Methodology, Resources, Supervision, Writing - review & editing. **Suzanne C. Purdy**: Conceptualization, Methodology, Project administration, Funding acquisition, Supervision, Writing - review & editing.

## CODE AVAILABILITY

No custom codes were used in any of the analyses.

## SUPPORTING INFORMATION

Additional supporting information may be found online in the supporting Information section at the end of this article.

## REFERENCES

Abraham, A., Pedregosa, F., Eickenberg, M., Gervais, P., Mueller, A., Kossaifi, J., Gramfort, A., Thirion, B., & Varoquaux, G. (2014). Machine learning for neuroimaging with scikit-learn. Frontiers in Neuroinformatics, 8, 14. https://doi.org/10.3389/fninf.2014.00014

Ahmmed, A. U., Ahmmed, A. A., Bath, J. R., Ferguson, M. A., Plack, C. J., & Moore, D. R. (2014). Assessment of children with suspected auditory processing disorder: a factor analysis study. Ear and Hearing, 35(3), 295–305. https://doi.org/10.1097/01.aud.0000441034.02052.0a

Alvand, A., Kuruvilla-Mathew, A., Kirk, I. J., Roberts, R. P., Pedersen, M., & Purdy, S. C. (2022). Altered brain network topology in children with auditory processing disorder: A resting-state multi-echo fMRI study. NeuroImage: Clinical, 35, 103139. https://doi.org/10.1016/j.nicl.2022.103139

Ameis, S. H., Fan, J., Rockel, C., Voineskos, A. N., Lobaugh, N. J., Soorya, L., Wang, A. T., Hollander, E., & Anagnostou, E. (2011). Impaired structural connectivity of socio- emotional circuits in autism spectrum disorders: a diffusion tensor imaging study. PloS One, 6(11), e28044. https://doi.org/10.1371/journal.pone.0028044

American Academy of Audiology (AAA). (2010). Guidelines for the diagnosis, treatment, and management of children and adults with central auditory processing disorder. https://audiology-web.s3.amazonaws.com/migrated/CAPD%20Guidelines%208-2010.pdf_539952af956c79.73897613.pdf

American Speech-Language-Hearing Association. (2005). *(Central) auditory processing disorders*. https://www.asha.org/content.aspx?id=10737450473

Andellini, M., Cannatà, V., Gazzellini, S., Bernardi, B., & Napolitano, A. (2015). Test-retest reliability of graph metrics of resting state MRI functional brain networks: A review. Journal of Neuroscience Methods, 253, 183–192. https://doi.org/10.1016/j.jneumeth.2015.05.020

Avants, B. B., Epstein, C. L., Grossman, M., & Gee, J. C. (2008). Symmetric diffeomorphic image registration with cross-correlation: evaluating automated labeling of elderly and neurodegenerative brain. Medical Image Analysis, 12(1), 26–41. https://doi.org/10.1016/j.media.2007.06.004

Bai, F., Shu, N., Yuan, Y., Shi, Y., Yu, H., Wu, D., Wang, J., Xia, M., He, Y., & Zhang, Z. (2012). Topologically convergent and divergent structural connectivity patterns between patients with remitted geriatric depression and amnestic mild cognitive impairment. The Journal of Neuroscience: The Official Journal of the Society for Neuroscience, 32(12), 4307–4318. https://doi.org/10.1523/JNEUROSCI.5061-11.2012

Baldi, S., Michielse, S., Vriend, C., van den Heuvel, M. P., van den Heuvel, O. A., Schruers, K. R. J., & Goossens, L. (2022). Abnormal white-matter rich-club organization in obsessive-compulsive disorder. Human Brain Mapping. https://doi.org/10.1002/hbm.25984

Barker, M. D., Kuruvilla-Mathew, A., & Purdy, S. C. (2017). Cortical Auditory-Evoked Potential and Behavioral Evidence for Differences in Auditory Processing between Good and Poor Readers. Journal of the American Academy of Audiology, 28(6), 534–545. https://doi.org/10.3766/jaaa.16054

Barkhof, F., Haller, S., & Rombouts, S. A. R. B. (2014). Resting-state functional MR imaging: a new window to the brain. Radiology, 272(1), 29–49. https://doi.org/10.1148/radiol.14132388

Bartel-Friedrich, S., Broecker, Y., Knoergen, M., & Koesling, S. (2010). Development of fMRI tests for children with central auditory processing disorders. In Vivo, 24(2), 201–209. https://www.ncbi.nlm.nih.gov/pubmed/20363995

Barulli, D., & Stern, Y. (2013). Efficiency, capacity, compensation, maintenance, plasticity: emerging concepts in cognitive reserve. Trends in Cognitive Sciences, 17(10), 502–509. https://doi.org/10.1016/j.tics.2013.08.012

Bassett, D. S., & Bullmore, E. T. (2009). Human brain networks in health and disease. Current Opinion in Neurology, 22(4), 340–347. https://doi.org/10.1097/WCO.0b013e32832d93dd

Bassett, D. S., & Sporns, O. (2017). Network neuroscience. Nature Neuroscience, 20(3), 353–364. https://doi.org/10.1038/nn.4502

Beare, R., Adamson, C., Bellgrove, M. A., Vilgis, V., Vance, A., Seal, M. L., & Silk, T. J. (2017). Altered structural connectivity in ADHD: a network based analysis. Brain Imaging and Behavior, 11(3), 846–858. https://doi.org/10.1007/s11682-016-9559-9

Beaulieu, C. (2002). The basis of anisotropic water diffusion in the nervous system - a technical review. NMR in Biomedicine, 15(7–8), 435–455. https://doi.org/10.1002/nbm.782

Belleau, E. L., Taubitz, L. E., & Larson, C. L. (2015). Imbalance of default mode and regulatory networks during externally focused processing in depression. Social Cognitive and Affective Neuroscience, 10(5), 744–751. https://doi.org/10.1093/scan/nsu117

Besser, J., Festen, J. M., Goverts, S. T., Kramer, S. E., & Pichora-Fuller, M. K. (2015). Speech-in-speech listening on the LiSN-S test by older adults with good audiograms depends on cognition and hearing acuity at high frequencies. Ear and Hearing, 36(1), 24–41. https://doi.org/10.1097/AUD.0000000000000096

Billeci, L., Calderoni, S., Tosetti, M., Catani, M., & Muratori, F. (2012). White matter connectivity in children with autism spectrum disorders: a tract-based spatial statistics study. BMC Neurology, 12, 148. https://doi.org/10.1186/1471-2377-12-148

Billingsley, R. L., McAndrews, M. P., Crawley, A. P., & Mikulis, D. J. (2001). Functional MRI of phonological and semantic processing in temporal lobe epilepsy. Brain: A Journal of Neurology, 124(Pt 6), 1218–1227. https://doi.org/10.1093/brain/124.6.1218

Buckner, R. L., Andrews-Hanna, J. R., & Schacter, D. L. (2008). The brain’s default network: anatomy, function, and relevance to disease. Annals of the New York Academy of Sciences, 1124, 1–38. https://doi.org/10.1196/annals.1440.011

Bullmore, E., & Sporns, O. (2009). Complex brain networks: graph theoretical analysis of structural and functional systems. Nature Reviews. Neuroscience, 10(3), 186–198. https://doi.org/10.1038/nrn2575

Cacace, A. T., & McFarland, D. J. (2013). Factors influencing tests of auditory processing: a perspective on current issues and relevant concerns. Journal of the American Academy of Audiology, 24(7), 572–589. https://doi.org/10.3766/jaaa.24.7.6

Cameron, Dillon, H., & Newall, P. (2006a). Development and evaluation of the listening in spatialized noise test. Ear and Hearing, 27(1), 30–42. https://doi.org/10.1097/01.aud.0000194510.57677.03

Cameron, Dillon, H., & Newall, P. (2006b). The listening in Spatialized Noise test: normative data for children. International Journal of Audiology, 45(2), 99–108. https://doi.org/10.1080/14992020500377931

Cameron, Glyde, H., & Dillon, H. (2011). Listening in Spatialized Noise—Sentences Test (LiSN-S): normative and retest reliability data for adolescents and adults up to 60 years of age. Journal of the American. https://www.ingentaconnect.com/contentone/aaa/jaaa/2011/00000022/00000010/art00007?crawler=true

Cameron, Sharon, & Dillon, H. (2007). Development of the Listening in Spatialized Noise-Sentences Test (LISN-S). Ear and Hearing, 28(2), 196–211. https://doi.org/10.1097/AUD.0b013e318031267f

Cameron, Sharon, & Dillon, H. (2008). The listening in spatialized noise-sentences test (LISN-S): comparison to the prototype LISN and results from children with either a suspected (central) auditory processing disorder or a confirmed language disorder. Journal of the American Academy of Audiology, 19(5), 377–391. https://doi.org/10.3766/jaaa.19.5.2

Cao, M., Wang, J.-H., Dai, Z.-J., Cao, X.-Y., Jiang, L.-L., Fan, F.-M., Song, X.-W., Xia, M.-R., Shu, N., Dong, Q., Milham, M. P., Castellanos, F. X., Zuo, X.-N., & He, Y. (2014). Topological organization of the human brain functional connectome across the lifespan. Developmental Cognitive Neuroscience, 7, 76–93. https://doi.org/10.1016/j.dcn.2013.11.004

Castellanos, F. X., Margulies, D. S., Kelly, C., Uddin, L. Q., Ghaffari, M., Kirsch, A., Shaw, D., Shehzad, Z., Di Martino, A., Biswal, B., Sonuga-Barke, E. J. S., Rotrosen, J., Adler, L. A., & Milham, M. P. (2008). Cingulate-Precuneus Interactions: A New Locus of Dysfunction in Adult Attention-Deficit/Hyperactivity Disorder. Biological Psychiatry, 63(3), 332–337. https://doi.org/10.1016/j.biopsych.2007.06.025

Chang, Y.-S., Owen, J. P., Desai, S. S., Hill, S. S., Arnett, A. B., Harris, J., Marco, E. J., & Mukherjee, P. (2014). Autism and sensory processing disorders: shared white matter disruption in sensory pathways but divergent connectivity in social-emotional pathways. PloS One, 9(7), e103038. https://doi.org/10.1371/journal.pone.0103038

Chow, H. M., Mar, R. A., Xu, Y., Liu, S., Wagage, S., & Braun, A. R. (2014). Embodied comprehension of stories: interactions between language regions and modality-specific neural systems. Journal of Cognitive Neuroscience, 26(2), 279–295. https://doi.org/10.1162/jocn_a_00487

Cieslak, M., Cook, P. A., He, X., Yeh, F.-C., Dhollander, T., Adebimpe, A., Aguirre, G. K., Bassett, D. S., Betzel, R. F., Bourque, J., Cabral, L. M., Davatzikos, C., Detre, J. A., Earl, E., Elliott, M. A., Fadnavis, S., Fair, D. A., Foran, W., Fotiadis, P., … Satterthwaite, T. D. (2021). QSIPrep: an integrative platform for preprocessing and reconstructing diffusion MRI data. Nature Methods, 1–4. https://doi.org/10.1038/s41592-021-01185-5

Colizza, V., Flammini, A., Serrano, M. A., & Vespignani, A. (2006). Detecting rich-club ordering in complex networks. Nature Physics, 2(2), 110–115. https://doi.org/10.1038/nphys209

Collin, G., Kahn, R. S., de Reus, M. A., Cahn, W., & van den Heuvel, M. P. (2014). Impaired rich club connectivity in unaffected siblings of schizophrenia patients. Schizophrenia Bulletin, 40(2), 438–448. https://doi.org/10.1093/schbul/sbt162

Collin, G., Sporns, O., Mandl, R. C. W., & van den Heuvel, M. P. (2014). Structural and functional aspects relating to cost and benefit of rich club organization in the human cerebral cortex. Cerebral Cortex, 24(9), 2258–2267. https://doi.org/10.1093/cercor/bht064

Cox, R. W., & Hyde, J. S. (1997). Software tools for analysis and visualization of fMRI data. NMR in Biomedicine, 10(4–5), 171–178. https://doi.org/10.1002/(sici)1099-1492(199706/08)10:4/5<171::aid-nbm453>3.0.co;2-l

Crossley, N. A., Mechelli, A., Scott, J., Carletti, F., Fox, P. T., McGuire, P., & Bullmore, E. T. (2014). The hubs of the human connectome are generally implicated in the anatomy of brain disorders. Brain: A Journal of Neurology, 137(Pt 8), 2382–2395. https://doi.org/10.1093/brain/awu132

Cui, W., Wang, S., Chen, B., & Fan, G. (2022). White matter structural network alterations in congenital bilateral profound sensorineural hearing loss children: A graph theory analysis. Hearing Research, 108521. https://doi.org/10.1016/j.heares.2022.108521

Daianu, M., Jahanshad, N., Nir, T. M., Jack, C. R., Jr, Weiner, M. W., Bernstein, M. A., Thompson, P. M., & Alzheimer’s Disease Neuroimaging Initiative. (2015). Rich club analysis in the Alzheimer’s disease connectome reveals a relatively undisturbed structural core network. Human Brain Mapping, 36(8), 3087–3103. https://doi.org/10.1002/hbm.22830

Daianu, M., Jahanshad, N., Villalon-Reina, J. E., Mendez, M. F., Bartzokis, G., Jimenez, E. E., Joshi, A., Barsuglia, J., & Thompson, P. M. (2014). Rich Club Network Analysis Shows Distinct Patterns of Disruption in Frontotemporal Dementia and Alzheimer’s Disease. Computational Diffusion MRI, 13–22. https://doi.org/10.1007/978-3-319-11182-7_2

Dawes, P., & Bishop, D. (2009). Auditory processing disorder in relation to developmental disorders of language, communication and attention: a review and critique. International Journal of Language & Communication Disorders / Royal College of Speech & Language Therapists, 44(4), 440–465. https://doi.org/10.1080/13682820902929073

Dawes, P., & Bishop, D. V. M. (2010). Psychometric profile of children with auditory processing disorder and children with dyslexia. Archives of Disease in Childhood, 95(6), 432–436. https://doi.org/10.1136/adc.2009.170118

Dawes, P., Bishop, D. V. M., Sirimanna, T., & Bamiou, D.-E. (2008). Profile and aetiology of children diagnosed with auditory processing disorder (APD). International Journal of Pediatric Otorhinolaryngology, 72(4), 483–489. https://doi.org/10.1016/j.ijporl.2007.12.007

Dawes, P., Sirimanna, T., Burton, M., Vanniasegaram, I., Tweedy, F., & Bishop, D. V. M. (2009). Temporal auditory and visual motion processing of children diagnosed with auditory processing disorder and dyslexia. Ear and Hearing, 30(6), 675–686. https://doi.org/10.1097/AUD.0b013e3181b34cc5

Delano-Wood, L., Stricker, N. H., Sorg, S. F., Nation, D. A., Jak, A. J., Woods, S. P., Libon, D. J., Delis, D. C., Frank, L. R., & Bondi, M. W. (2012). Posterior cingulum white matter disruption and its associations with verbal memory and stroke risk in mild cognitive impairment. Journal of Alzheimer’s Disease: JAD, 29(3), 589–603. https://doi.org/10.3233/JAD-2012-102103

DeSalvo, M. N., Douw, L., Tanaka, N., Reinsberger, C., & Stufflebeam, S. M. (2014). Altered structural connectome in temporal lobe epilepsy. Radiology, 270(3), 842–848. https://doi.org/10.1148/radiol.13131044

Dhollander, T., Clemente, A., Singh, M., Boonstra, F., Civier, O., Duque, J. D., Egorova, N., Enticott, P., Fuelscher, I., Gajamange, S., Genc, S., Gottlieb, E., Hyde, C., Imms, P., Kelly, C., Kirkovski, M., Kolbe, S., Liang, X., Malhotra, A., … Caeyenberghs, K. (2021). Fixel-based Analysis of Diffusion MRI: Methods, Applications, Challenges and Opportunities. NeuroImage, 241, 118417. https://doi.org/10.1016/j.neuroimage.2021.118417

Dillon, H., & Cameron, S. (2021). Separating the Causes of Listening Difficulties in Children. Ear and Hearing, 42(5), 1097–1108. https://doi.org/10.1097/AUD.0000000000001069

Dillon, H., Cameron, S., Glyde, H., Wilson, W., & Tomlin, D. (2012). An opinion on the assessment of people who may have an auditory processing disorder. Journal of the American Academy of Audiology, 23(2), 97–105. https://doi.org/10.3766/jaaa.23.2.4

Ercan, E. S., Suren, S., Bacanlı, A., Yazıcı, K. U., Callı, C., Ardic, U. A., Aygunes, D., Kosova, B., Ozyurt, O., Aydın, C., & Rohde, L. A. (2016). Altered structural connectivity is related to attention deficit/hyperactivity subtypes: A DTI study. Psychiatry Research. Neuroimaging, 256, 57–64. https://doi.org/10.1016/j.pscychresns.2016.04.002

Esteban, O., Markiewicz, C. J., Burns, C., Goncalves, M., Jarecka, D., Ziegler, E., Berleant, S., Ellis, D. G., Pinsard, B., Madison, C., Waskom, M., Notter, M. P., Clark, D., Manhães-Savio, A., Clark, D., Jordan, K., Dayan, M., Halchenko, Y. O., Loney, F., … Ghosh, S. (2022). nipy/nipype: 1.7.1. https://doi.org/10.5281/zenodo.6415183

Fan, L., Li, H., Zhuo, J., Zhang, Y., Wang, J., Chen, L., Yang, Z., Chu, C., Xie, S., Laird, A. R., Fox, P. T., Eickhoff, S. B., Yu, C., & Jiang, T. (2016). The Human Brainnetome Atlas: A New Brain Atlas Based on Connectional Architecture. Cerebral Cortex, 26(8), 3508–3526. https://doi.org/10.1093/cercor/bhw157

Fang, H., Wu, Q., Li, Y., Ren, Y., Li, C., Xiao, X., Xiao, T., Chu, K., & Ke, X. (2020). Structural networks in children with autism spectrum disorder with regression: A graph theory study. Behavioural Brain Research, 378, 112262. https://doi.org/10.1016/j.bbr.2019.112262

Farah, R., Schmithorst, V. J., Keith, R. W., & Holland, S. K. (2014). Altered white matter microstructure underlies listening difficulties in children suspected of auditory processing disorders: a DTI study. Brain and Behavior, 4(4), 531–543. https://doi.org/10.1002/brb3.237

Ferstl, E. C., Neumann, J., Bogler, C., & von Cramon, D. Y. (2008). The extended language network: a meta-analysis of neuroimaging studies on text comprehension. Human Brain Mapping, 29(5), 581–593. https://doi.org/10.1002/hbm.20422

Ferstl, E. C., Rinck, M., & von Cramon, D. Y. (2005). Emotional and temporal aspects of situation model processing during text comprehension: an event-related fMRI study. Journal of Cognitive Neuroscience, 17(5), 724–739. https://doi.org/10.1162/0898929053747658

Fonov, V. S., Evans, A. C., McKinstry, R. C., Almli, C. R., & Collins, D. L. (2009). Unbiased nonlinear average age-appropriate brain templates from birth to adulthood. NeuroImage, 47, S102. https://doi.org/10.1016/S1053-8119(09)70884-5

Fornito, A., Zalesky, A., & Breakspear, M. (2015). The connectomics of brain disorders. Nature Reviews. Neuroscience, 16(3), 159–172. https://doi.org/10.1038/nrn3901

Fornito, A., Zalesky, A., & Bullmore, E. T. (Eds.). (2016a). Chapter 4 - Node Degree and Strength. In Fundamentals of Brain Network Analysis (pp. 115–136). Academic Press. https://doi.org/10.1016/B978-0-12-407908-3.00004-2

Fornito, A., Zalesky, A., & Bullmore, E. T. (Eds.). (2016b). Chapter 5 - Centrality and Hubs. In Fundamentals of Brain Network Analysis (pp. 137–161). Academic Press. https://doi.org/10.1016/B978-0-12-407908-3.00005-4

Fornito, A., Zalesky, A., & Bullmore, E. T. (Eds.). (2016c). Chapter 6 - Components, Cores, and Clubs. In Fundamentals of Brain Network Analysis (pp. 163–206). Academic Press. https://doi.org/10.1016/B978-0-12-407908-3.00006-6

Fornito, A., Zalesky, A., & Bullmore, E. T. (Eds.). (2016d). Chapter 7 - Paths, Diffusion, and Navigation. In Fundamentals of Brain Network Analysis (pp. 207–255). Academic Press. https://doi.org/10.1016/B978-0-12-407908-3.00007-8

Frost, J. A., Binder, J. R., Springer, J. A., Hammeke, T. A., Bellgowan, P. S., Rao, S. M., & Cox, R. W. (1999). Language processing is strongly left lateralized in both sexes. Evidence from functional MRI. Brain: A Journal of Neurology, 122 ( *Pt 2*), 199–208. https://doi.org/10.1093/brain/122.2.199

Garyfallidis, E., Brett, M., Amirbekian, B., Rokem, A., van der Walt, S., Descoteaux, M., Nimmo-Smith, I., & Dipy Contributors. (2014). Dipy, a library for the analysis of diffusion MRI data. Frontiers in Neuroinformatics, 8, 8. https://doi.org/10.3389/fninf.2014.00008

Gitelman, D. R., Nobre, A. C., Sonty, S., Parrish, T. B., & Mesulam, M.-M. (2005). Language network specializations: an analysis with parallel task designs and functional magnetic resonance imaging. NeuroImage, 26(4), 975–985. https://doi.org/10.1016/j.neuroimage.2005.03.014

Gokula, R., Sharma, M., Cupples, L., & Valderrama, J. T. (2019). Comorbidity of Auditory Processing, Attention, and Memory in Children With Word Reading Difficulties. Frontiers in Psychology, 10, 2383. https://doi.org/10.3389/fpsyg.2019.02383

Gorgolewski, Burns, C. D., Madison, C., Clark, D., Halchenko, Y. O., Waskom, M. L., & Ghosh, S. S. (2011). Nipype: a flexible, lightweight and extensible neuroimaging data processing framework in python. Frontiers in Neuroinformatics, 5, 13. https://doi.org/10.3389/fninf.2011.00013

Gorgolewski, K. J., Alfaro-Almagro, F., Auer, T., Bellec, P., Capotă, M., Chakravarty, M. M., Churchill, N. W., Cohen, A. L., Craddock, R. C., Devenyi, G. A., Eklund, A., Esteban, O., Flandin, G., Ghosh, S. S., Guntupalli, J. S., Jenkinson, M., Keshavan, A., Kiar, G., Liem, F., … Poldrack, R. A. (2017). BIDS apps: Improving ease of use, accessibility, and reproducibility of neuroimaging data analysis methods. PLoS Computational Biology, 13(3), e1005209. https://doi.org/10.1371/journal.pcbi.1005209

Goya-Maldonado, R., Brodmann, K., Keil, M., Trost, S., Dechent, P., & Gruber, O. (2016). Differentiating unipolar and bipolar depression by alterations in large-scale brain networks. Human Brain Mapping, 37(2), 808–818. https://doi.org/10.1002/hbm.23070

Hagmann, P., Cammoun, L., Gigandet, X., Meuli, R., Honey, C. J., Wedeen, V. J., & Sporns, O. (2008). Mapping the structural core of human cerebral cortex. PLoS Biology, 6(7), e159. https://doi.org/10.1371/journal.pbio.0060159

Halliday, L. F., Tuomainen, O., & Rosen, S. (2017). Auditory processing deficits are sometimes necessary and sometimes sufficient for language difficulties in children: Evidence from mild to moderate sensorineural hearing loss. Cognition, 166, 139–151. https://doi.org/10.1016/j.cognition.2017.04.014

Hind, S. E., Haines-Bazrafshan, R., Benton, C. L., Brassington, W., Towle, B., & Moore, D. R. (2011). Prevalence of clinical referrals having hearing thresholds within normal limits. International Journal of Audiology, 50(10), 708–716. https://doi.org/10.3109/14992027.2011.582049

Huang, L., Zheng, W., Wu, C., Wei, X., Wu, X., Wang, Y., & Zheng, H. (2015). Diffusion Tensor Imaging of the Auditory Neural Pathway for Clinical Outcome of Cochlear Implantation in Pediatric Congenital Sensorineural Hearing Loss Patients. PloS One, 10(10), e0140643. https://doi.org/10.1371/journal.pone.0140643

Hugdahl, K. (2011). Fifty years of dichotic listening research - still going and going and…. Brain and Cognition, 76(2), 211–213. https://doi.org/10.1016/j.bandc.2011.03.006

Iliadou, V. V., Chermak, G. D., Bamiou, D.-E., Rawool, V. W., Ptok, M., Purdy, S., Jutras, B., Moncrieff, D., Stokkereit Mattsson, T., Ferre, J. M., Fox, C., Grech, H., Geffner, D., Hedjever, M., Bellis, T. J., Nimatoudis, I., Eleftheriadis, N., Pedersen, E. R., Weihing, J., … Musiek, F. E. (2018). Letter to the Editor: An Affront to Scientific Inquiry Re: Moore, D. R. (2018) Editorial: Auditory Processing Disorder, Ear Hear, 39, 617-620 [Review of *Letter to the Editor: An Affront to Scientific Inquiry Re: Moore, D. R. (2018) Editorial: Auditory Processing Disorder, Ear Hear, 39, 617-620*]. Ear and Hearing, 39(6), 1236–1242. https://doi.org/10.1097/AUD.0000000000000644

Kaiser, R. H., Andrews-Hanna, J. R., Wager, T. D., & Pizzagalli, D. A. (2015). Large-Scale Network Dysfunction in Major Depressive Disorder: A Meta-analysis of Resting-State Functional Connectivity. JAMA Psychiatry, 72(6), 603–611. https://doi.org/10.1001/jamapsychiatry.2015.0071

Kambara, T., Brown, E. C., Jeong, J.-W., Ofen, N., Nakai, Y., & Asano, E. (2017). Spatio- temporal dynamics of working memory maintenance and scanning of verbal information. Clinical Neurophysiology: Official Journal of the International Federation of Clinical Neurophysiology, 128(6), 882–891. https://doi.org/10.1016/j.clinph.2017.03.005

Keith, W., Purdy, S., Baily, M. R., & Kay, F. M. (2019). *New Zealand guidelines on auditory processing disorder*. https://researchspace.auckland.ac.nz/handle/2292/48927

Keown, C. L., Datko, M. C., Chen, C. P., Maximo, J. O., Jahedi, A., & Müller, R.-A. (2017). Network Organization Is Globally Atypical in Autism: A Graph Theory Study of Intrinsic Functional Connectivity. Biological Psychiatry: Cognitive Neuroscience and Neuroimaging, 2(1), 66–75. https://doi.org/10.1016/j.bpsc.2016.07.008

Li, D., Liu, W., Yan, T., Cui, X., Zhang, Z., Wei, J., Ma, Y., Zhang, N., Xiang, J., & Wang, B. (2020). Disrupted Rich Club Organization of Hemispheric White Matter Networks in Bipolar Disorder. Frontiers in Neuroinformatics, 14, 39. https://doi.org/10.3389/fninf.2020.00039

Li, W., Li, J., Wang, J., Zhou, P., Wang, Z., Xian, J., & He, H. (2016). Functional Reorganizations of Brain Network in Prelingually Deaf Adolescents. Neural Plasticity, 2016, 9849087. https://doi.org/10.1155/2016/9849087

Li, Z., Zhu, Q., Geng, Z., Song, Z., Wang, L., & Wang, Y. (2015). Study of functional connectivity in patients with sensorineural hearing loss by using resting-state fMRI. International Journal of Clinical and Experimental Medicine, 8(1), 569–578. https://www.ncbi.nlm.nih.gov/pubmed/25785031

Liu, T., Yan, Y., Ai, J., Chen, D., Wu, J., Fang, B., & Yan, T. (2021). Disrupted rich-club organization of brain structural networks in Parkinson’s disease. Brain Structure & Function. https://doi.org/10.1007/s00429-021-02319-3

Lou, C., Cross, A. M., Peters, L., Ansari, D., & Joanisse, M. F. (2021). Rich-club structure contributes to individual variance of reading skills via feeder connections in children with reading disabilities. Developmental Cognitive Neuroscience, 49, 100957. https://doi.org/10.1016/j.dcn.2021.100957

Lu, Y., Li, Y., Feng, Q., Shen, R., Zhu, H., Zhou, H., & Zhao, Z. (2021). Rich-Club Analysis of the Structural Brain Network in Cases with Cerebral Small Vessel Disease and Depression Symptoms. Cerebrovascular Diseases, 1–10. https://doi.org/10.1159/000517243

Maier-Hein, K. H., Neher, P. F., Houde, J.-C., Côté, M.-A., Garyfallidis, E., Zhong, J., Chamberland, M., Yeh, F.-C., Lin, Y.-C., Ji, Q., Reddick, W. E., Glass, J. O., Chen, D. Q., Feng, Y., Gao, C., Wu, Y., Ma, J., He, R., Li, Q., … Descoteaux, M. (2017). The challenge of mapping the human connectome based on diffusion tractography. Nature Communications, 8(1), 1349. https://doi.org/10.1038/s41467-017-01285-x

Maslov, S., & Sneppen, K. (2002). Specificity and stability in topology of protein networks. Science, 296(5569), 910–913. https://doi.org/10.1126/science.1065103

McAuley, J. J., da Fontoura Costa, L., & Caetano, T. S. (2007). Rich-club phenomenon across complex network hierarchies. Applied Physics Letters, 91(8), 084103. https://doi.org/10.1063/1.2773951

McFarland, D. J., & Cacace, A. T. (2014). Modality specificity is the preferred method for diagnosing the auditory processing disorder (APD): response to Moore and Ferguson [Review of *Modality specificity is the preferred method for diagnosing the auditory processing disorder (APD): response to Moore and Ferguson*]. Journal of the American Academy of Audiology, 25(7), 698–699. neurotechcenter.org. https://www.ncbi.nlm.nih.gov/pubmed/25365373

Mealings, K., & Cameron, S. (2019). Investigating Auditory Spectral and Temporal Resolution Deficits in Children with Reading Difficulties. Journal of the American Academy of Audiology, 30(6), 533–543. https://doi.org/10.3766/jaaa.17142

Merlet, S. L., & Deriche, R. (2013). Continuous diffusion signal, EAP and ODF estimation via Compressive Sensing in diffusion MRI. Medical Image Analysis, 17(5), 556–572. https://doi.org/10.1016/j.media.2013.02.010

Meunier, D., Achard, S., Morcom, A., & Bullmore, E. (2009). Age-related changes in modular organization of human brain functional networks. NeuroImage, 44(3), 715–723. https://doi.org/10.1016/j.neuroimage.2008.09.062

Moncrieff, D. W. (2011). Dichotic listening in children: age-related changes in direction and magnitude of ear advantage. Brain and Cognition, 76(2), 316–322. https://doi.org/10.1016/j.bandc.2011.03.013

Moore, D. R. (2012). Listening difficulties in children: bottom-up and top-down contributions. Journal of Communication Disorders, 45(6), 411–418. https://doi.org/10.1016/j.jcomdis.2012.06.006

Moore, D. R. (2018). Editorial: Auditory Processing Disorder. Ear and Hearing, 39(4), 617–620. https://doi.org/10.1097/AUD.0000000000000582

Moore, D. R., & Hunter, L. L. (2013). Auditory processing disorder (APD) in children: A marker of neurodevelopmental syndrome. Hearing, Balance and Communication, 11(3), 160–167. https://doi.org/10.3109/21695717.2013.821756

Mori, S., Crain, B. J., Chacko, V. P., & van Zijl, P. C. (1999). Three-dimensional tracking of axonal projections in the brain by magnetic resonance imaging. Annals of Neurology, 45(2), 265–269. https://doi.org/10.1002/1531-8249(199902)45:2<265::aid-ana21>3.0.co;2-3

Mori, Susumu, & van Zijl, P. C. M. (2002). Fiber tracking: principles and strategies - a technical review. NMR in Biomedicine, 15(7–8), 468–480. https://doi.org/10.1002/nbm.781

Nakai, Y., Sugiura, A., Brown, E. C., Sonoda, M., Jeong, J.-W., Rothermel, R., Luat, A. F., Sood, S., & Asano, E. (2019). Four-dimensional functional cortical maps of visual and auditory language: Intracranial recording. Epilepsia, 60(2), 255–267. https://doi.org/10.1111/epi.14648

Noppeney, U., Friston, K. J., & Price, C. J. (2004). Degenerate neuronal systems sustaining cognitive functions. Journal of Anatomy, 205(6), 433–442. https://doi.org/10.1111/j.0021-8782.2004.00343.x

O’Connor, K. (2012). Auditory processing in autism spectrum disorder: a review. Neuroscience and Biobehavioral Reviews, 36(2), 836–854. https://doi.org/10.1016/j.neubiorev.2011.11.008

Oldham, S., Ball, G., & Fornito, A. (2021). Early and late development of hub connectivity in the human brain. Current Opinion in Psychology. https://doi.org/10.1016/j.copsyc.2021.10.010

Opsahl, T., Colizza, V., Panzarasa, P., & Ramasco, J. J. (2008). Prominence and control: the weighted rich-club effect. Physical Review Letters, 101(16), 168702. https://doi.org/10.1103/PhysRevLett.101.168702

Owen, J. P., Marco, E. J., Desai, S., Fourie, E., Harris, J., Hill, S. S., Arnett, A. B., & Mukherjee, P. (2013). Abnormal white matter microstructure in children with sensory processing disorders. NeuroImage. Clinical, 2, 844–853. https://doi.org/10.1016/j.nicl.2013.06.009

Park, K. H., Chung, W.-H., Kwon, H., & Lee, J.-M. (2018). Evaluation of Cerebral White Matter in Prelingually Deaf Children Using Diffusion Tensor Imaging. BioMed Research International, 2018, 6795397. https://doi.org/10.1155/2018/6795397

Pedersen, M., & Omidvarnia, A. (2016). Further Insight into the Brain’s Rich-Club Architecture. The Journal of Neuroscience: The Official Journal of the Society for Neuroscience, 36(21), 5675–5676. https://doi.org/10.1523/JNEUROSCI.0754-16.2016

Peng, Z., Yang, X., Xu, C., Wu, X., Yang, Q., Wei, Z., Zhou, Z., Verguts, T., & Chen, Q. (2021). Aberrant rich club organization in patients with obsessive-compulsive disorder and their unaffected first-degree relatives. NeuroImage. Clinical, 32, 102808. https://doi.org/10.1016/j.nicl.2021.102808

Pluta, A., Wolak, T., Czajka, N., Lewandowska, M., Cieśla, K., Rusiniak, M., Grudzień, D., & Skarżyński, H. (2014). Reduced resting-state brain activity in the default mode network in children with (central) auditory processing disorders. Behavioral and Brain Functions: BBF, 10(1), 33. https://doi.org/10.1186/1744-9081-10-33

Ponton, C. W., Moore, J. K., & Eggermont, J. J. (1996). Auditory brain stem response generation by parallel pathways: differential maturation of axonal conduction time and synaptic transmission. Ear and Hearing, 17(5), 402–410. https://doi.org/10.1097/00003446-199610000-00006

Purdy, S. C., Sharma, M., & Morgan, A. (2018). Measuring Perceptions of Classroom Listening in Typically Developing Children and Children with Auditory Difficulties Using the LIFE-UK Questionnaire. Journal of the American Academy of Audiology, 29(7), 656–667. https://doi.org/10.3766/jaaa.17053

Qin, P., Wu, X., Huang, Z., Duncan, N. W., Tang, W., Wolff, A., Hu, J., Gao, L., Jin, Y., Wu, X., Zhang, J., Lu, L., Wu, C., Qu, X., Mao, Y., Weng, X., Zhang, J., & Northoff, G. (2015). How are different neural networks related to consciousness? Annals of Neurology, 78(4), 594–605. https://doi.org/10.1002/ana.24479

Raffelt, D. A., Smith, R. E., Ridgway, G. R., Tournier, J.-D., Vaughan, D. N., Rose, S., Henderson, R., & Connelly, A. (2015). Connectivity-based fixel enhancement: Whole-brain statistical analysis of diffusion MRI measures in the presence of crossing fibres. NeuroImage, 117, 40–55. https://doi.org/10.1016/j.neuroimage.2015.05.039

Raffelt, D. A., Tournier, J.-D., Smith, R. E., Vaughan, D. N., Jackson, G., Ridgway, G. R., & Connelly, A. (2017). Investigating white matter fibre density and morphology using fixel-based analysis. NeuroImage, 144(Pt A), 58–73. https://doi.org/10.1016/j.neuroimage.2016.09.029

Raichle, M. E., MacLeod, A. M., Snyder, A. Z., Powers, W. J., Gusnard, D. A., & Shulman, G. L. (2001). A default mode of brain function. Proceedings of the National Academy of Sciences of the United States of America, 98(2), 676–682. https://doi.org/10.1073/pnas.98.2.676

Ray, S., Miller, M., Karalunas, S., Robertson, C., Grayson, D. S., Cary, R. P., Hawkey, E., Painter, J. G., Kriz, D., Fombonne, E., Nigg, J. T., & Fair, D. A. (2014). Structural and functional connectivity of the human brain in autism spectrum disorders and attention-deficit/hyperactivity disorder: A rich club-organization study. Human Brain Mapping, 35(12), 6032–6048. https://doi.org/10.1002/hbm.22603

Rocca, R., Coventry, K. R., Tylén, K., Staib, M., Lund, T. E., & Wallentin, M. (2020). Language beyond the language system: Dorsal visuospatial pathways support processing of demonstratives and spatial language during naturalistic fast fMRI. NeuroImage, 216, 116128. https://doi.org/10.1016/j.neuroimage.2019.116128

Roger, E., Pichat, C., Torlay, L., David, O., Renard, F., Banjac, S., Attyé, A., Minotti, L., Lamalle, L., Kahane, P., & Baciu, M. (2020). Hubs disruption in mesial temporal lobe epilepsy. A resting-state fMRI study on a language-and-memory network. Human Brain Mapping, 41(3), 779–796. https://doi.org/10.1002/hbm.24839

Rorden, C., Karnath, H.-O., & Bonilha, L. (2007). Improving Lesion-Symptom Mapping. Journal of Cognitive Neuroscience, 19(7), 1081–1088. https://doi.org/10.1162/jocn.2007.19.7.1081

Roup, C. M., Wiley, T. L., Safady, S. H., & Stoppenbach, D. T. (1998). Tympanometric Screening Norms for Adults. American Journal of Audiology, 7(2), 55–60. https://doi.org/10.1044/1059-0889(1998/014)

Rubinov, M., & Bullmore, E. (2013). Schizophrenia and abnormal brain network hubs. Dialogues in Clinical Neuroscience, 15(3), 339–349. https://doi.org/10.31887/dcns.2013.15.3/mrubinov

Rubinov, M., & Sporns, O. (2010). Complex network measures of brain connectivity: uses and interpretations. NeuroImage, 52(3), 1059–1069. https://doi.org/10.1016/j.neuroimage.2009.10.003

Ruytjens, L., Albers, F., van Dijk, P., Wit, H., & Willemsen, A. (2006). Neural responses to silent lipreading in normal hearing male and female subjects. The European Journal of Neuroscience, 24(6), 1835–1844. https://doi.org/10.1111/j.1460-9568.2006.05072.x

Sa de Almeida, J., Meskaldji, D.-E., Loukas, S., Lordier, L., Gui, L., Lazeyras, F., & Hüppi, P. S. (2021). Preterm birth leads to impaired rich-club organization and fronto-paralimbic/limbic structural connectivity in newborns. NeuroImage, 225, 117440. https://doi.org/10.1016/j.neuroimage.2020.117440

Santesso, D. L., Meuret, A. E., Hofmann, S. G., Mueller, E. M., Ratner, K. G., Roesch, E. B., & Pizzagalli, D. A. (2008). Electrophysiological correlates of spatial orienting towards angry faces: a source localization study. Neuropsychologia, 46(5), 1338–1348. https://doi.org/10.1016/j.neuropsychologia.2007.12.013

Schmidt, S. A., Akrofi, K., Carpenter-Thompson, J. R., & Husain, F. T. (2013). Default mode, dorsal attention and auditory resting state networks exhibit differential functional connectivity in tinnitus and hearing loss. PloS One, 8(10), e76488. https://doi.org/10.1371/journal.pone.0076488

Schmithorst, V. J., Farah, R., & Keith, R. W. (2013). Left ear advantage in speech-related dichotic listening is not specific to auditory processing disorder in children: A machine-learning fMRI and DTI study. NeuroImage. Clinical, 3, 8–17. https://doi.org/10.1016/j.nicl.2013.06.016

Schmithorst, V. J., Holland, S. K., & Plante, E. (2011). Diffusion tensor imaging reveals white matter microstructure correlations with auditory processing ability. Ear and Hearing, 32(2), 156–167. https://doi.org/10.1097/AUD.0b013e3181f7a481

Sharma, M., Purdy, S. C., & Kelly, A. S. (2009). Comorbidity of auditory processing, language, and reading disorders. Journal of Speech, Language, and Hearing Research: JSLHR, 52(3), 706–722. https://doi.org/10.1044/1092-4388(2008/07-0226)

Sharma, M., Purdy, S. C., & Kelly, A. S. (2014). The Contribution of Speech-Evoked Cortical Auditory Evoked Potentials to the Diagnosis and Measurement of Intervention Outcomes in Children with Auditory Processing Disorder. Seminars in Hearing, 35(01), 051–064. https://doi.org/10.1055/s-0033-1363524

Shu, N., Duan, Y., Huang, J., Ren, Z., Liu, Z., Dong, H., Barkhof, F., Li, K., & Liu, Y. (2018). Progressive brain rich-club network disruption from clinically isolated syndrome towards multiple sclerosis. NeuroImage. Clinical, 19, 232–239. https://doi.org/10.1016/j.nicl.2018.03.034

Sihvonen, A. J., Virtala, P., Thiede, A., Laasonen, M., & Kujala, T. (2021). Structural white matter connectometry of reading and dyslexia. NeuroImage, 241, 118411. https://doi.org/10.1016/j.neuroimage.2021.118411

Soares, J. M., Marques, P., Alves, V., & Sousa, N. (2013). A hitchhiker’s guide to diffusion tensor imaging. Frontiers in Neuroscience, 7, 31. https://doi.org/10.3389/fnins.2013.00031

Sporns, O. (2011). The human connectome: a complex network. Annals of the New York Academy of Sciences, 1224, 109–125. https://doi.org/10.1111/j.1749-6632.2010.05888.x

Stam, C. J. (2014). Modern network science of neurological disorders. Nature Reviews. Neuroscience, 15(10), 683–695. https://doi.org/10.1038/nrn3801

Stewart, H. J., Cash, E. K., Hunter, L. L., Maloney, T., Vannest, J., & Moore, D. R. (2022). Speech cortical activation and connectivity in typically developing children and those with listening difficulties. NeuroImage. Clinical, 36(103172), 103172. https://doi.org/10.1016/j.nicl.2022.103172

Termenon, M., Jaillard, A., Delon-Martin, C., & Achard, S. (2016). Reliability of graph analysis of resting state fMRI using test-retest dataset from the Human Connectome Project. NeuroImage, 142, 172–187. https://doi.org/10.1016/j.neuroimage.2016.05.062

Tononi, G., Sporns, O., & Edelman, G. M. (1999). Measures of degeneracy and redundancy in biological networks. Proceedings of the National Academy of Sciences of the United States of America, 96(6), 3257–3262. https://doi.org/10.1073/pnas.96.6.3257

Torgerson, C. M., Irimia, A., Goh, S. Y. M., & Van Horn, J. D. (2015). The DTI connectivity of the human claustrum. Human Brain Mapping, 36(3), 827–838. https://doi.org/10.1002/hbm.22667

Tournier, J.-D., Smith, R., Raffelt, D., Tabbara, R., Dhollander, T., Pietsch, M., Christiaens, D., Jeurissen, B., Yeh, C.-H., & Connelly, A. (2019). MRtrix3: A fast, flexible and open software framework for medical image processing and visualisation. NeuroImage, 202, 116137. https://doi.org/10.1016/j.neuroimage.2019.116137

Tuch, D. S. (2004). Q-ball imaging. Magnetic Resonance in Medicine: Official Journal of the Society of Magnetic Resonance in Medicine / Society of Magnetic Resonance in Medicine, 52(6), 1358–1372. https://doi.org/10.1002/mrm.20279

Tustison, N. J., Avants, B. B., Cook, P. A., Zheng, Y., Egan, A., Yushkevich, P. A., & Gee, J. C. (2010). N4ITK: improved N3 bias correction. IEEE Transactions on Medical Imaging, 29(6), 1310–1320. https://doi.org/10.1109/TMI.2010.2046908

Valenzuela, M. J., Breakspear, M., & Sachdev, P. (2007). Complex mental activity and the aging brain: molecular, cellular and cortical network mechanisms. Brain Research Reviews, 56(1), 198–213. https://doi.org/10.1016/j.brainresrev.2007.07.007

van den Heuvel, M. P., Mandl, R. C. W., Stam, C. J., Kahn, R. S., & Hulshoff Pol, H. E. (2010). Aberrant frontal and temporal complex network structure in schizophrenia: a graph theoretical analysis. The Journal of Neuroscience: The Official Journal of the Society for Neuroscience, 30(47), 15915–15926. https://doi.org/10.1523/JNEUROSCI.2874-10.2010

van den Heuvel, M. P., & Sporns, O. (2011). Rich-club organization of the human connectome. The Journal of Neuroscience: The Official Journal of the Society for Neuroscience, 31(44), 15775–15786. https://doi.org/10.1523/JNEUROSCI.3539-11.2011

van den Heuvel, M. P., & Sporns, O. (2013). Network hubs in the human brain. Trends in Cognitive Sciences, 17(12), 683–696. https://doi.org/10.1016/j.tics.2013.09.012

Van Den Heuvel, M. P., Sporns, O., & Collin, G. (2013). Abnormal rich club organization and functional brain dynamics in schizophrenia. JAMA: The Journal of the American Medical Association. https://jamanetwork.com/journals/jamapsychiatry/article-abstract/1695592

Veraart, J., Novikov, D. S., Christiaens, D., Ades-Aron, B., Sijbers, J., & Fieremans, E. (2016). Denoising of diffusion MRI using random matrix theory. NeuroImage, 142, 394–406. https://doi.org/10.1016/j.neuroimage.2016.08.016

Vogelzang, M., Thiel, C. M., Rosemann, S., Rieger, J. W., & Ruigendijk, E. (2021). Effects of age-related hearing loss and hearing aid experience on sentence processing. Scientific Reports, 11(1), 5994. https://doi.org/10.1038/s41598-021-85349-5

Wang, B., Wang, G., Wang, X., Cao, R., Xiang, J., Yan, T., Li, H., Yoshimura, S., Toichi, M., & Zhao, S. (2021). Rich-Club Analysis in Adults With ADHD Connectomes Reveals an Abnormal Structural Core Network. Journal of Attention Disorders, 25(8), 1068– 1079. https://doi.org/10.1177/1087054719883031

Wang, Y., Deng, F., Jia, Y., Wang, J., Zhong, S., Huang, H., Chen, L., Chen, G., Hu, H., Huang, L., & Huang, R. (2019). Disrupted rich club organization and structural brain connectome in unmedicated bipolar disorder. Psychological Medicine, 49(3), 510–518. https://doi.org/10.1017/S0033291718001150

Wang, Y., Zhong, S., Chen, G., Liu, T., Zhao, L., Sun, Y., Jia, Y., & Huang, L. (2018). Altered cerebellar functional connectivity in remitted bipolar disorder: A resting-state functional magnetic resonance imaging study. The Australian and New Zealand Journal of Psychiatry, 52(10), 962–971. https://doi.org/10.1177/0004867417745996

Wilson, W. J. (2018). Evolving the concept of APD. International Journal of Audiology, 57(4), 240–248. https://doi.org/10.1080/14992027.2017.1409438

Winkler, A. M., Ridgway, G. R., Webster, M. A., Smith, S. M., & Nichols, T. E. (2014). Permutation inference for the general linear model. NeuroImage, 92, 381–397. https://doi.org/10.1016/j.neuroimage.2014.01.060

Xia, M., Wang, J., & He, Y. (2013). BrainNet Viewer: a network visualization tool for human brain connectomics. PloS One, 8(7), e68910. https://doi.org/10.1371/journal.pone.0068910

Xie, X., Liu, Y., Han, X., Liu, P., Qiu, H., Li, J., & Yu, H. (2019). Differences in Intrinsic Brain Abnormalities Between Patients With Left- and Right-Sided Long-Term Hearing Impairment. Frontiers in Neuroscience, 13, 206. https://doi.org/10.3389/fnins.2019.00206

Xue, C., Sun, H., Hu, G., Qi, W., Yue, Y., Rao, J., Yang, W., Xiao, C., & Chen, J. (2020). Disrupted Patterns of Rich-Club and Diverse-Club Organizations in Subjective Cognitive Decline and Amnestic Mild Cognitive Impairment. Frontiers in Neuroscience, 14, 575652. https://doi.org/10.3389/fnins.2020.575652

Yang, M., Chen, H.-J., Liu, B., Huang, Z.-C., Feng, Y., Li, J., Chen, J.-Y., Zhang, L.-L., Ji, H., Feng, X., Zhu, X., & Teng, G.-J. (2014). Brain structural and functional alterations in patients with unilateral hearing loss. Hearing Research, 316, 37–43. https://doi.org/10.1016/j.heares.2014.07.006

Yarkoni, T., Poldrack, R. A., Nichols, T. E., Van Essen, D. C., & Wager, T. D. (2011). Large-scale automated synthesis of human functional neuroimaging data. Nature Methods, 8(8), 665–670. https://doi.org/10.1038/nmeth.1635

Yeh, F.-C., Verstynen, T. D., Wang, Y., Fernández-Miranda, J. C., & Tseng, W.-Y. I. (2013). Deterministic diffusion fiber tracking improved by quantitative anisotropy. PloS One, 8(11), e80713. https://doi.org/10.1371/journal.pone.0080713

Yeh, F.-C., Wedeen, V. J., & Tseng, W.-Y. I. (2010). Generalized q-sampling imaging. IEEE Transactions on Medical Imaging, 29(9), 1626–1635. https://doi.org/10.1109/TMI.2010.2045126

Yeh, F.-C., Zaydan, I. M., Suski, V. R., Lacomis, D., Richardson, R. M., Maroon, J. C., & Barrios-Martinez, J. (2019). Differential tractography as a track-based biomarker for neuronal injury. NeuroImage, 202, 116131. https://doi.org/10.1016/j.neuroimage.2019.116131

Yuan, W., Wade, S. L., & Babcock, L. (2015). Structural connectivity abnormality in children with acute mild traumatic brain injury using graph theoretical analysis. Human Brain Mapping, 36(2), 779–792. https://doi.org/10.1002/hbm.22664

Zalesky, A., Fornito, A., & Bullmore, E. T. (2010). Network-based statistic: identifying differences in brain networks. NeuroImage, 53(4), 1197–1207. https://doi.org/10.1016/j.neuroimage.2010.06.041

Zalesky, A., Fornito, A., Cocchi, L., Gollo, L. L., van den Heuvel, M. P., & Breakspear, M. (2016). Connectome sensitivity or specificity: which is more important? NeuroImage, 142, 407–420. https://doi.org/10.1016/j.neuroimage.2016.06.035

Zalesky, A., Fornito, A., Harding, I. H., Cocchi, L., Yücel, M., Pantelis, C., & Bullmore, E. T. (2010). Whole-brain anatomical networks: does the choice of nodes matter? NeuroImage, 50(3), 970–983. https://doi.org/10.1016/j.neuroimage.2009.12.027

Zhang, Y., Brady, M., & Smith, S. (2001). Segmentation of brain MR images through a hidden Markov random field model and the expectation-maximization algorithm. IEEE Transactions on Medical Imaging, 20(1), 45–57. https://doi.org/10.1109/42.906424

Zhou, S., & Mondragon, R. J. (2004). The rich-club phenomenon in the Internet topology. IEEE Communications Letters, 8(3), 180–182. https://doi.org/10.1109/LCOMM.2004.823426

