## Supplementary Information for "Altered structural connectome of children with Auditory Processing Disorder: A diffusion MRI study"

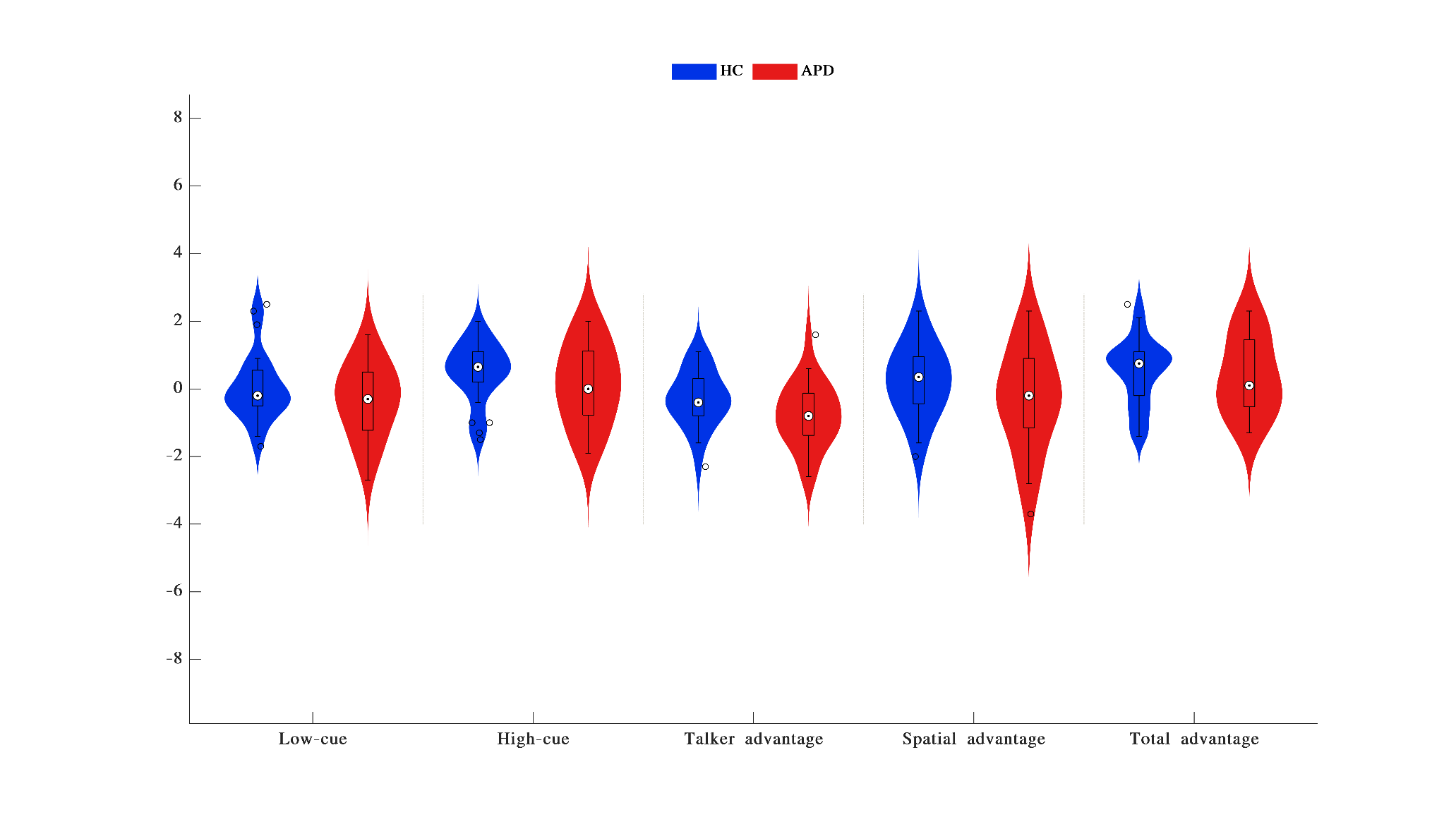
**Figure S1** Distribution of LiSN-S z-scores (Low-cue, High-cue, Talker advantage, Spatial advantage, Total advantage) for APD and HC participants. HC – healthy control, APD – auditory processing disorder, LiSN-S - listening-in-spatialized-noise-sentences.

**Table S1**

*Results from between-group differences based on FA and GFA connectivity measures*

| **Connectivity** | | **ROI** | ***p* value** | ***t* stat** | **Bonferroni correction** |
| --- | --- | --- | --- | --- | --- |
| FA | 144 | | 0.0003 | 3.9764 | 0.0653 |
|  | 64 | | 0.0001 | 3.9805 | 0.0644 |
| GFA | 144 | | 0.0003 | 3.9048 | 0.0827 |
|  | 64 | | 0.0001 | 3.8764 | 0.0907 |

***Note:*** ROI – region of interest, *t* stat – test statistic, FA – fractional anisotropy, GFA – generalized fractional anisotropy.
